# Enhanced power and transferability for genetics-driven metabolomic biomarker discovery in admixed American cohorts

**DOI:** 10.64898/2026.09.03.26362043

**Authors:** Tin Orešković, Danyao Jin, Eirini Trichia, Diego Aguilar-Ramirez, Yu Xu, Carles Foguet, Tabitha Hubbard, Michael Turner, Alejandra Vergara-Lope, Luisa Fernández-Chirino, Laurent Gil, Michael R. Hill, Jesús Alegre-Díaz, Jaime Berumen, Sarah Lewington, Jonathan R. Emberson, Pablo Kuri-Morales, Roberto Tapia-Conyer, Jason M. Torres, Michael Inouye

## Abstract

Despite metabolomics transforming our understanding of risk factors and aetiology of metabolic diseases, profiling is rarely performed for people of non-European ancestries, on whom much of metabolic disease burden falls. Metabolome-wide association studies (MWAS) can be performed using genetic scores to predict metabolomic traits, helping address these inequities; however, their performance in populations of admixed American (AMR) ancestries is unexplored. We evaluated 141 genetic scores, developed in an INTERVAL Study sample of European (EUR) genetic ancestries, in the Mexico City Prospective Study (MCPS; *n*=132,336), obtaining a median predictive *R*² of 0.027. Training Bayesian ridge models within MCPS substantially improved performance, with a median *R*² of 0.083 on a withheld 20% subset. MCPS-trained models also outperformed INTERVAL-trained models among UK Biobank participants of AMR ancestries (*n*=600; median *R*^2^: 0.070 vs. 0.046). Finally, among AMR participants of the All of Us cohort, using MCPS-trained (vs. INTERVAL-trained) models to predict metabolomic traits yielded five times as many significant associations (FDR-corrected *P*<0.05) across three cardiometabolic diseases: ischaemic heart disease, type 2 diabetes, and chronic kidney disease. The genetic scores are openly available at the OmicsPred portal (www.OmicsPred.org), enabling better-powered analyses in diverse AMR cohorts and helping reduce global inequities in omics research.

## Introduction

The use of multi-omic data has improved the breadth and depth of our understanding of the aetiologies of common diseases^1–5^. However, as collecting omic data is costly, time consuming, and logistically complex, large-scale cohorts often include such data only on a subset of participants and a narrow range of traits, or not at all, which limits the scope and statistical power of analyses^6^.

The obstacles to collecting multi-omic data disproportionately affect populations in non-European and resource-constrained settings, limiting both participation in biomedical research and access to its benefits^7,8^. Among the groups most acutely underrepresented in multi-omics research are various Latin American populations, despite their bearing a large and growing burden of disease attributable to obesity and cardiometabolic diseases^9–11^. According to the Global Burden of Disease Study 2023, high body mass index, fasting plasma glucose, and systolic blood pressure were the three risk factors with the largest attributable burden of disease in each region of the “Latin America and the Caribbean” super region (Andean, Tropical, and Central Latin America, as well as the Caribbean)^10^. In Mexico, for instance, type 2 diabetes is more common, more poorly controlled, and has a much larger effect on premature mortality than in high-income countries^12,13^. Furthermore, certain genetic variants strongly associated with type 2 diabetes in several populations with admixed American (AMR) genetic ancestries^14–16^ are rare or absent in European populations. Indigenous American (IAM) genetic ancestries proportion (compared with, for example, the European [EUR] ancestries proportion among individuals of AMR ancestries) and related measures have also been associated with type 2 diabetes and body mass index in Mexico and the United States, associations that may also depend on the urban environment and its social context^17–19^. Together, these observations underscore the need for multi-omics research in AMR populations, particularly on metabolic disease biomarkers and disease aetiology.

Since genetic data are increasingly widely available and more affordable to collect, approaches to transcriptome-, proteome-, and metabolome-wide association studies (TWAS, PWAS, MWAS, respectively) that rely exclusively on genetic models to impute omic traits can help address the difficulties of conducting multi-omics research in underrepresented populations^3,20–24^. Previous work has demonstrated the wide applicability of genetic prediction models across omic modalities and in relation to several diseases^25^. Unsurprisingly, however, it has also reported that there is an attenuation of prediction accuracy in non-European populations, as existing models have been trained on samples of EUR genetic ancestries^26,27^.

Given the large burden of obesity and cardiometabolic disease in AMR populations, we focus here on addressing inequities in multi-omics research by using existing nuclear magnetic resonance (NMR) spectroscopy metabolomics data in the Mexico City Prospective Study (MCPS) (**Extended Data Fig. 1**)^21,28^. Using MCPS data, we first quantify the extent of attenuation in the performance of existing genetic prediction models for metabolomic traits, trained in a sample of EUR ancestries. Next, we develop new genetic prediction models for metabolomics in MCPS that address this attenuation, then validate their performance internally and in two external samples of individuals of AMR ancestries outside Mexico. We then conduct an MWAS using the newly developed genetic prediction models in a USA-based sample of AMR ancestries, demonstrating substantially increased power to identify metabolomic biomarkers for three cardiometabolic diseases. To enable and enhance multi-omics research in diverse cohorts of AMR ancestries in Mexico, Latin America, and beyond, we make these new genetic models for metabolomic traits freely available through OmicsPred^29^ (https://www.omicspred.org/).

## Methods

### Genetic and NMR metabolomic data from the Mexico City Prospective Study

The Mexico City Prospective Study (MCPS) is a prospective cohort of more than 150,000 participants of age 35 or older when they were recruited between 1998 and 2004 from two contiguous districts (Coyoacán and Iztapalapa) in Mexico City. The design and aims of the study have been described previously^30^. Briefly, as part of the baseline assessment, the recruitment teams visited individual households and collected information on the participants’ sociodemographic and lifestyle characteristics, medical history and medication use, performed various physical measurements, and collected EDTA blood samples^30^.

DNA was extracted from buffy-coat samples and genotyped using an Illumina Global Screening Array (GSA) v.2 beadchip, as described previously in detail^28^. Genotype imputation was performed using the Trans-Omics for Precision Medicine (TOPMed) reference panel^31^, and the final dataset of 140,829 participants after sample- and variant-level quality control, mapped to genome build GRCh38. Per-individual AMR and EUR (as well as “African” and “East Asian”) genetic ancestries proportions used in this work were first estimated using the ADMIXTURE program^32^ in a combined dataset of 1,000 samples from unrelated MCPS participants, 2,248 samples from the 1000 Genomes and Human Genome Diversity Project^33^ reference panel, and 716 samples from the Metabolic Syndrome in Indigenous Sample (MAIS) cohort^34,35^. The remaining MCPS samples were then projected onto the admixture model to obtain estimates for the complete cohort.

The EDTA plasma samples have been used to perform nuclear magnetic resonance (NMR) spectroscopy–based metabolomic traits profiling using the Nightingale Health Plc. platform, which generates spectra from which 225 measures are quantified as absolute concentrations or ratios. The details of the Nightingale Health NMR platform in general, including its standardised quality control procedures, have been described previously in detail, in general and as used in the MCPS^36,37^. This work, like that of Xu et al.^25^, developing the set of genetic scores in the INTERVAL Study^38^ described below, is focused only on the 141 directly measured metabolomic traits, excluding those derived from other traits (**Supplementary Table 1**).

### Evaluation of EUR-trained models for NMR metabolomic traits in the Mexico City Prospective Study

Xu et al.^25^ developed genetic scores for over 17,000 traits across several omic modalities using Bayesian ridge models trained on data from EUR-ancestries participants of the INTERVAL Study^38^, a cohort of UK blood donors. For the 141 metabolomic traits measured directly from INTERVAL participants’ serum samples via the Nightingale Health NMR platform, the genetic variants (aligned to the GRCh38 genome build) used as variables were selected based on genome-wide statistical significance from a genome-wide association study (GWAS) of each of the traits in a sample of 37,359 participants. Four fifths of this sample were then used in the training of Bayesian ridge models and the remaining 20% withheld for internal validation of the resulting scores. In addition to the withheld subset of the INTERVAL sample, the models were also validated in an external sample of EUR genetic ancestries (from the UK Biobank)^1^, as well as Chinese-, Indian- and Malay-ancestries subsets of the Singapore Multi-Ethnic Cohort (MEC)^39^. The genetic scores were released through OmicsPred^29^ (https://www.omicspred.org/).

We applied the genetic scores for the 141 Nightingale Health NMR metabolomic traits to MCPS participants, using the Polygenic Score Catalog (PGSC) Calculator^40^, and evaluated their performance by comparing the predictions with the measured metabolomic trait values (the variant matching rates for all 141 genetic scores were above the default threshold of 75% and are reported in **Supplementary Table 1**). Before the performance evaluation, we performed quality control and pre-processing in a manner similar to the steps employed by Xu et al. in INTERVAL^25^.

Of the 140,829 MCPS participants with imputed genetic data, 138,524 also had metabolomic data. Although the vast majority of MCPS participants are of AMR genetic ancestries^28^, we performed additional ancestry classification to identify participants of AMR ancestries according to the same criteria that we used in external cohorts, described below. Briefly, as part of the PGSC Calculator pipeline with its default parameters^40,41^, we ran a principal component analysis (PCA) on the Human Genome Diversity Project (HGDP) + 1000 Genomes Project (1kGP) reference panel^33^ and projected the MCPS samples onto the resulting PC space. We then estimated the probability of each MCPS participant’s membership of each continental-level ancestry group in the reference panel using a 20-nearest-neighbour algorithm. The probability of having AMR genetic ancestry was estimated to be ≥ 90% for 137,844 participants, who were retained in the sample, while 680 participants with estimated probability below 90% were excluded. A further 5,508 participants with ≥ 30% of metabolomic data missing were removed, resulting in a dataset of 132,336 participants. As missingness varied across the traits, the trait-specific sample sizes are reported in **Supplementary Table 1**.

We natural log transformed the metabolomic trait values (after replacing 0 values with a randomised value between 0.001 and 0.9 × the lowest observed non-zero value for the trait) and regressed them on age, sex, district, the month of donation, hour of donation, metabolomic processing duration, and the first seven genetic PCs. The first seven PCs were used as covariates instead of the typically used 10 because, in MCPS, this was the maximal set of consecutive PCs with normally distributed loadings, while non-normally distributed loadings observed for subsequent PCs are indicative of long-range linkage disequilibrium (LD)^28,42^. The residual values from regressions of the log-transformed values on the selected covariates were finally inverse-rank normalised. The predicted values were similarly adjusted for age, sex, district, and the first seven genetic PCs, and inverse-rank normalised. We assessed the predictive performance of the scores in terms of variance explained (*R*^2^) between the values predicted by the genetic score and the measured trait (after the transformations and adjustments described above).

In secondary analyses, to provide evidence on whether, as expected^26^, genetic ancestry contributed to an attenuation of the overall performance observed in the present validation in MCPS compared to in previously reported external validations in samples of EUR ancestries, we assessed the performance of the models in subsets of MCPS defined by higher IAM (≥ 70%) or EUR (≥ 70%) ancestries proportions (n.b. all participants in these subsets are of AMR genetic ancestries, with ≥ 90% probability, as described above). We further assessed the performance in subsets defined by: sex; age groups (35–49, 50–64, and ≥ 65 years); and baseline health status, defined by the presence or absence of self-reported diabetes or glycosylated haemoglobin (HbA_1c_) > 6.5 at baseline, as well as by the presence or absence of diabetes and a range of other diseases about which the participants were also asked at baseline (self-reported cardiovascular disease, cancer, chronic kidney disease, chronic obstructive pulmonary disease, peptic ulcer, and liver cirrhosis).

### Training and evaluation of new models in the Mexico City Prospective Study

Following the evaluation of the predictive performance of INTERVAL-trained models in MCPS, we trained new models for the prediction of the 141 directly measured Nightingale Health NMR metabolomic traits on data from the MCPS. Mirroring the methods employed by Xu et al.^25^ in INTERVAL, we selected genetic variants with genome-wide significant associations with each metabolomic trait (*P* < 5 × 10^−8^) from GWAS previously conducted using MCPS samples of 123,022 to 130,841 participants (summary statistics shared via private communication; GWAS details are briefly described in in **Supplementary Note 1**). We also performed a thinning step at an *r*^2^ threshold of 0.8 to remove LD dependencies among the variants, applied an MAF threshold of 0.5%, and excluded multi-allelic and ambiguous variants, as was done for the original set of models trained in INTERVAL.

To train new models for each of the 141 traits, we used the genome-wide significantly associated genetic variants and randomly selected subsets of 80% of the up to 132,336 MCPS participants that we used in the evaluation of the equivalent INTERVAL-trained models (training sample sizes varied across traits due to differences in missingness, and are reported in **Supplementary Table 2**; median *n* = 105,868). The remaining 20% of the data were used for internal validation of the models (minimal *n* = 24,879, median and maximal *n* = 26,468). While the use of the general NMR metabolomics MCPS sample for the GWAS used to identify genetic variants may cause concern about a possible leakage of information from the validation sample into the training sample, this practice has not been found to lead to poor generalisation of Bayesian ridge models trained in INTERVAL to an external sample of EUR ancestries, the UKB^25^; the newly trained models have similarly been externally validated in samples of AMR genetic ancestries, as described below.

We used Bayesian ridge models (matching the method used by Xu et al.^25^) because of their computational efficiency, making them suitable for the large number of target traits, as well as their predictive performance, as previously assessed for omics traits^25,43^. We set the two prior gamma distribution hyperparameters for the models to the non-informative *α*_1_ = *α*_2_ = *λ*_1_ = *λ_2_* = 10^−5^. This approach has previously been observed to yield near-optimal performance while avoiding a computationally costly per-trait fine-tuning of the hyperparameters^25,44^. Nevertheless, to empirically validate this choice with metabolomic data in this context, we randomly selected 10 of the 141 traits and performed a hyperparameter optimisation experiment using only the training data (80% of the total sample) in a five-fold cross validation scheme. We used a random search procedure, which is more efficient than an exhaustive grid search and yields similar results^45^. We evaluated a search space of 200 combinations, sampling the four hyperparameters from a log-uniform distribution between 10^−10^ and 10^10^, in addition to a run with the non-informative priors. The Bayesian ridge models with non-informative priors indeed performed nearly as well as with the optimised set of hyperparameters for all 10 traits, as measured by the *R*^2^ averaged across the five withheld folds in the cross validation scheme (**Supplementary Table 3**).

We assessed the predictive performance of the new models in withheld subsets of the sample not used in training (20% of the data) in terms of *R*^2^ and compared it to that of the INTERVAL-trained models in the same withheld subsets; we used median *R*^2^ to compare the overall performance across the 141 traits, as well as the slope (*λ*) of the line of best fit through the origin between the MCPS-trained and INTERVAL-trained models’ *R*^2^ metrics. As above, we further assessed the performance in subsets of the withheld 20% defined in the same manner: split by genetic ancestry proportion, sex, and health status at baseline.

### External validation in the UK Biobank

The UK Biobank (UKB) is a large-scale prospective cohort of approximately 500,000 individuals recruited from across the UK between 2006 and 2010 of ages 37–73 at the baseline assessment. Metabolomic data in the UKB were generated from plasma samples using the same Nightingale Health NMR platform used in MCPS^25^ which, along with the imputed genetic data in the UKB^46^, enabled an evaluation of the performance of all 141 new MCPS-trained models and a comparison to that of the INTERVAL-trained models.

We identified a subset of 613 UKB participants of AMR genetic ancestries using the same HGDP + 1kGP reference panel and ancestry classification algorithm as described above for MCPS (requiring again a probability of AMR ancestries ≥ 90%). We transformed the metabolomic trait values as in MCPS, and adjusted them for age, sex, recruitment centre, the month of donation, hour of donation, metabolomic processing duration, and the first ten genetic PCs. There were up to 600 participants with complete genetic, metabolomic, and covariate data (trait-specific sample sizes are reported in **Supplementary Table 4**).

After mapping the variants used in the MCPS- and INTERVAL-trained models from genome build GRCh38 to GRCh37 using the LiftOver program^47^, we predicted the 141 metabolomic traits via the PGSC Calculator^40^ for the same set of AMR participants of the UKB. The variant matching rate was above the PGSC Calculator default threshold of 75% for all 141 traits for both sets of models. We adjusted the predicted values and transformed them as in MCPS. We compared the predictive performance of the MCPS-trained models to that of INTERVAL-trained models in terms of the median *R*^2^ between their predictions and the measured metabolomic traits, as well as the slope *λ*, as in internal validation. We further assessed the performance of the models in subsets of UKB sample split by sex and age group categories (37–49, 50–64, and ≥ 65 years).

### External validation in the All of Us Research Program

The All of Us (AoU) Research Program is a longitudinal cohort study aiming to enrol one million or more participants across the United States that started enrolment in 2018 with a focus on populations historically underrepresented in biomedical research. The design and genomic data of the AoU cohort have been described previously in detail^48,49^. Briefly, AoU collects a wide range of health-related information, including electronic health records (EHR) and survey responses, alongside biospecimens used to generate large-scale whole-genome sequencing (WGS) data for 414,840 participants aligned to the GRCh38 genome build.

To futher externally validate a subset of the newly developed MCPS-trained models and compare their performance to that of INTERVAL-trained models in another sample of AMR genetic ancestries, we used WGS and EHR data from AoU participants. There were 64,155 participants with estimated probability of AMR genetic ancestry ≥ 90% and with WGS data passing standard genomic quality control procedures^48^. The AoU Program Genomics Investigators identified pariticipants of AMR genetic ancestries based on an ancestry inference pipeline involving a random forest classifier trained on the same HGDP and 1kGP reference panel^33^ that we used in MCPS and the UKB.

We defined six metabolomic traits in the AoU EHR data matched to traits measured by the Nightingale Health NMR spectroscopy platform in the MCPS and UKB cohorts: HDL cholesterol, LDL cholesterol, VLDL cholesterol, total triglycerides, glucose, and creatinine. We defined these traits based on records of clinical measurements using the Observational Medical Outcomes Partnership (OMOP) concept IDs, which map to Logical Observation Identifiers Names and Codes (LOINC), an international standard ontology for health measurements, observations, and documents^50^. Specifically, we used concepts 3007070, 3053286, and 4195503 for HDL cholesterol; concepts 3009966, 3028288, 3028437, 3035899, and 3053341 for LDL cholesterol; concepts 3007352 and 3009596 for VLDL cholesterol; concept 3022192 for triglycerides; concept 3004501 for glucose; and concept 3016723 for creatinine. We also predicted the six traits for AoU participants of AMR ancestries using both the new MCPS-trained models and the INTERVAL-trained models, again via the PGSC Calculator^40^.

We restricted the data to measurements with explicitly stated units (such as mg/dL, mmol/l) and removed observations with values above the 99th percentile, as some of these were data-entry errors or artifacts (e.g. “10000000”). We then harmonised the values to mmol/l units to match those of the Nightingale NMR platform. To align the age profile more closely with that of the MCPS, we excluded measurements taken when the participant had under 35 years of age, and retained only the earliest record among the remaining measurements. Finally, we removed any participants for whom the trait could not be predicted using the MCPS- and INTERVAL-trained models due to genetic data availability, or for whom any of the covariates used to adjust the values (listed below) were missing. Following these filtering steps, the trait-specific sample sizes within the AoU AMR subset ranged from 4,349 participants for VLDL cholesterol to 21,737 participants for glucose (**Supplementary Table 5**).

We log transformed the measured AoU metabolomic traits as in MCPS and adjusted them for age at measurement, sex, and the first ten genetic PCs. We then inverse-rank normalised the residual values from these regressions. We also adjusted the predicted trait values for age, sex, and the first ten genetic PCs, and inverse-rank normalised them. We compared the predictive performance of the MCPS-trained and INTERVAL-trained models in terms of the mean *R*^2^.

### Predicted metabolome–wide association analyses in All of Us

To evaluate the utility of ancestry-matched genetic prediction in downstream epidemiological analyses, we assessed associations between genetically predicted metabolomic traits and three high-burden cardiometabolic diseases: ischaemic heart disease (4,387 cases and 24,592 controls), type 2 diabetes (10,633 cases and 18,346 controls), and chronic kidney disease (3,552 cases and 25,427 controls).

The analyses were restricted to 28,979 AoU participants of AMR genetic ancestries who were of age 35 years or older at the time of their last recorded EHR event. We extracted longitudinal ICD-10 and ICD-9 code events mapped to clinical phenotypes matching the three diseases using the PhecodeX system (**Supplementary Table 6**), an expansion of the standard phecode framework designed to better reflect the granularity and organisation of ICD-10 codes^51^.

We predicted the metabolomic traits using both sets of models via the PGSC Calculator^40^: the analyses were performed for 139 of the 141 traits because, for the remaining two (acetate and pyruvate), less than 75% of the variants used in the models were present in AoU data, the default threshold in the PGSC Calculator workflow (the overall variant matching rate for both sets of models is reported in **Supplementary Table 7**).

We inverse-rank normalised the genetic scores and included the age at the last available EHR event, sex, and the first ten genetic principal components as covariates in logistic regression models to estimate the relationships between the odds of disease and the predicted metabolomic traits. We compared the two sets of predicted traits in terms of the number of their statistically significant associations with the diseases, defined using the Benjamini– Hochberg false discovery rate (FDR)^52^ at a threshold of 0.05 to adjust for multiple testing, as well as the Pearson’s correlation *r* between the log odds estimates.

### Ethics

Our work complies with the relevant ethical regulations: all studies used in this work were approved by the relevant board or committee. The Mexico City Prospective Study was approved by the Mexican Ministry of Health, the Mexican National Council for Science and Technology (0595 P-M), the Central Oxford Research Ethics Committee (C99.260), and the Medical Ethics Committee of the National Autonomous University of Mexico (FMED/CEI/MHU/2020); all MCPS participants provided written informed consent. The UK Biobank has approval from the North West Multi-centre Research Ethics Committee (reference 21/NW/0157), which is renewed every five years; all UKB participants provided informed consent. The All of Us Research Program protocol and participant-facing materials were reviewed by its Institutional Review Board, which follows the regulations and guidance of the NIH Office for Human Research Protections for all studies, ensuring that the rights and welfare of research participants are overseen and protected uniformly; all AoU participants provided informed consent in person or electronically.

## Results

### Evaluation of EUR-trained models for NMR metabolomic traits in the Mexico City Prospective Study

We evaluated the performance of existing models^25,29^ for 141 NMR metabolomic traits trained in INTERVAL Study^38^ using data from MCPS participants of AMR ancestries with genetic, metabolomic, and covariate data (**Methods**). In MCPS, the INTERVAL-trained NMR metabolomic models had a median *R*^2^ of 0.027 (interquartile range, IQR: 0.016; **Fig. 1** and **Supplementary Table 1**). In contrast, the performance of INTERVAL-trained metabolomic models in external validation on data from individuals of EUR ancestries (median *R*^2^: 0.067; IQR: 0.039)^53^ was substantially greater, consistent with a sensitivity of the genetic models for metabolomics to ancestry. Notably, the performance of these models in MCPS was lower than in external validation in cohorts of other non-European ancestries (**Fig. 1**), suggesting the challenge of model transferability is particularly acute for populations of admixed American ancestries.

**Fig. 1:**
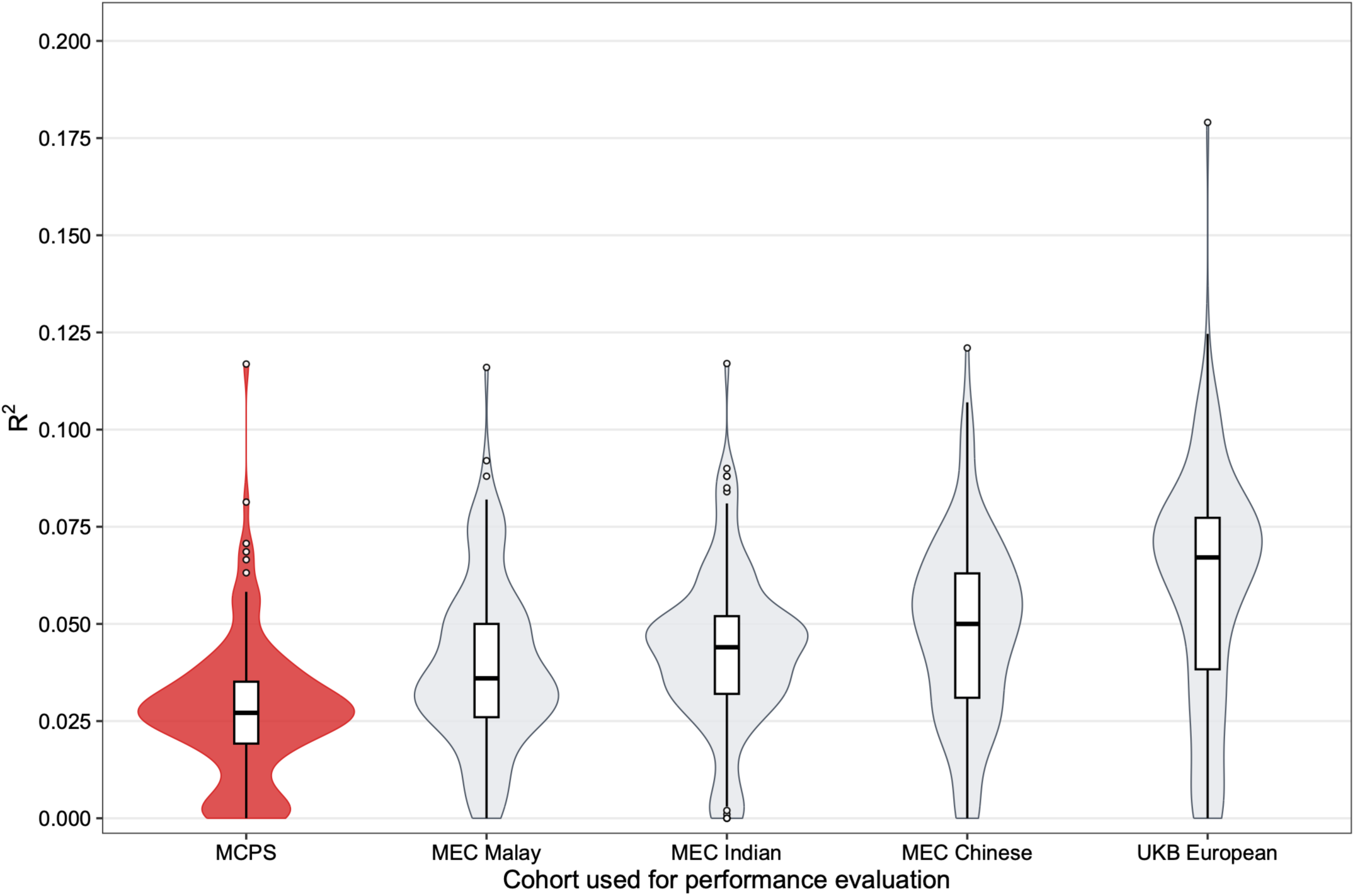
Performance of INTERVAL-trained models in MCPS and other external validation cohorts. The boxplots show the median, interquartile ranges (IQR), 1.5 times the ranges, and outliers in the predictive performance in terms of variance explained (*R*^2^) of INTERVAL Study–trained models for 141 metabolomic traits in cohorts external to INTERVAL. The violin plots behind the boxplots show the kernel density estimates of the *R*^2^ of the same models. The cohorts are the Mexico Prospective Cohort Study (MCPS; median *R*^2^: 0.027; IQR: 0.024) and, for context, other cohorts external to INTERVAL where the performance of the models has been previously assessed: the subsets of the Singapore Multi-Ethnic Cohort (MEC) of Malay (MA; median *R*²: 0.036; IQR: 0.024), Indian (IN; median *R*²: 0.044; IQR: 0.020), and Chinese (CN; median *R*²: 0.050; IQR: 0.032) ancestries, as well as the subset of the UK Biobank (UKB) of European (EUR) ancestries (median *R*²: 0.067; IQR: 0.039).

To test this, we assessed INTERVAL-trained metabolomic models in the 62,830 MCPS participants with a higher (≥ 70%) estimated IAM genetic ancestries proportion and found performance to be further attenuated (median *R*^2^: 0.022; IQR: 0.013; **Fig. 2**). Conversely, for the up to 891 MCPS participants with a higher (≥ 70%) estimated EUR genetic ancestries proportion, INTERVAL-trained metabolomic model performance was improved (median *R*^2^: 0.038; IQR: 0.023).

**Fig. 2:**
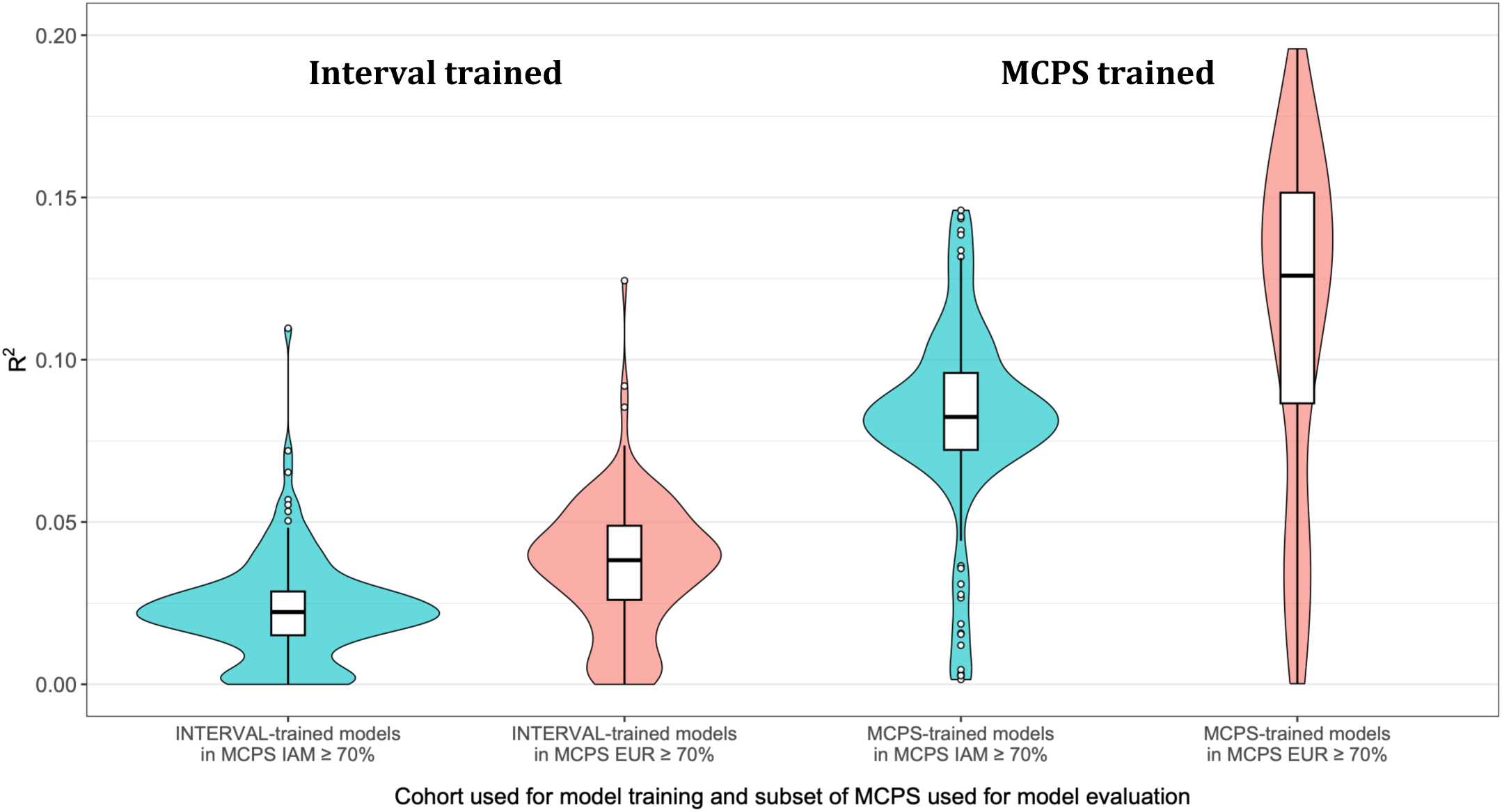
Performance of MCPS- and INTERVAL-trained models among MCPS participants with higher Indigenous American and higher European genetic ancestries proportions. Predictive performance of 141 INTERVAL Study–trained models and MCPS-trained models for metabolomic traits in subsets of the Mexico City Prospective Study (MCPS) defined by higher Indigenous American (IAM) ancestries proportion (≥ 70%) and higher European (EUR) genetic ancestries proportion (≥ 70%). MCPS-trained models were trained in 80% of the MCPS sample and evaluated on the remaining 20%, with the latter subset stratified by the same genetic ancestry proportion categories as the overall sample for the evaluation of the INTERVAL-trained models.

### Training and evaluation of new models in the Mexico City Prospective Study

In an 80% subset of MCPS participants with available data, we trained models for the 141 NMR metabolomic traits using Bayesian ridge regression on genetic variants selected through genome-wide association studies conducted in MCPS (**Methods**). The performance of the new models was evaluated in a withheld 20% subset of MCPS (∼26,000 participants, depending on trait). The MCPS-trained metabolomic models had substantially higher performance on the withheld MCPS subset (median *R*^2^: 0.083 and IQR: 0.021) than the INTERVAL-trained models (median *R*^2^: 0.027 and IQR: 0.016), a more than three-fold increase in variance explained per metabolomic trait (**Fig. 3a**; **Supplementary Table 2**).

**Fig. 3:**
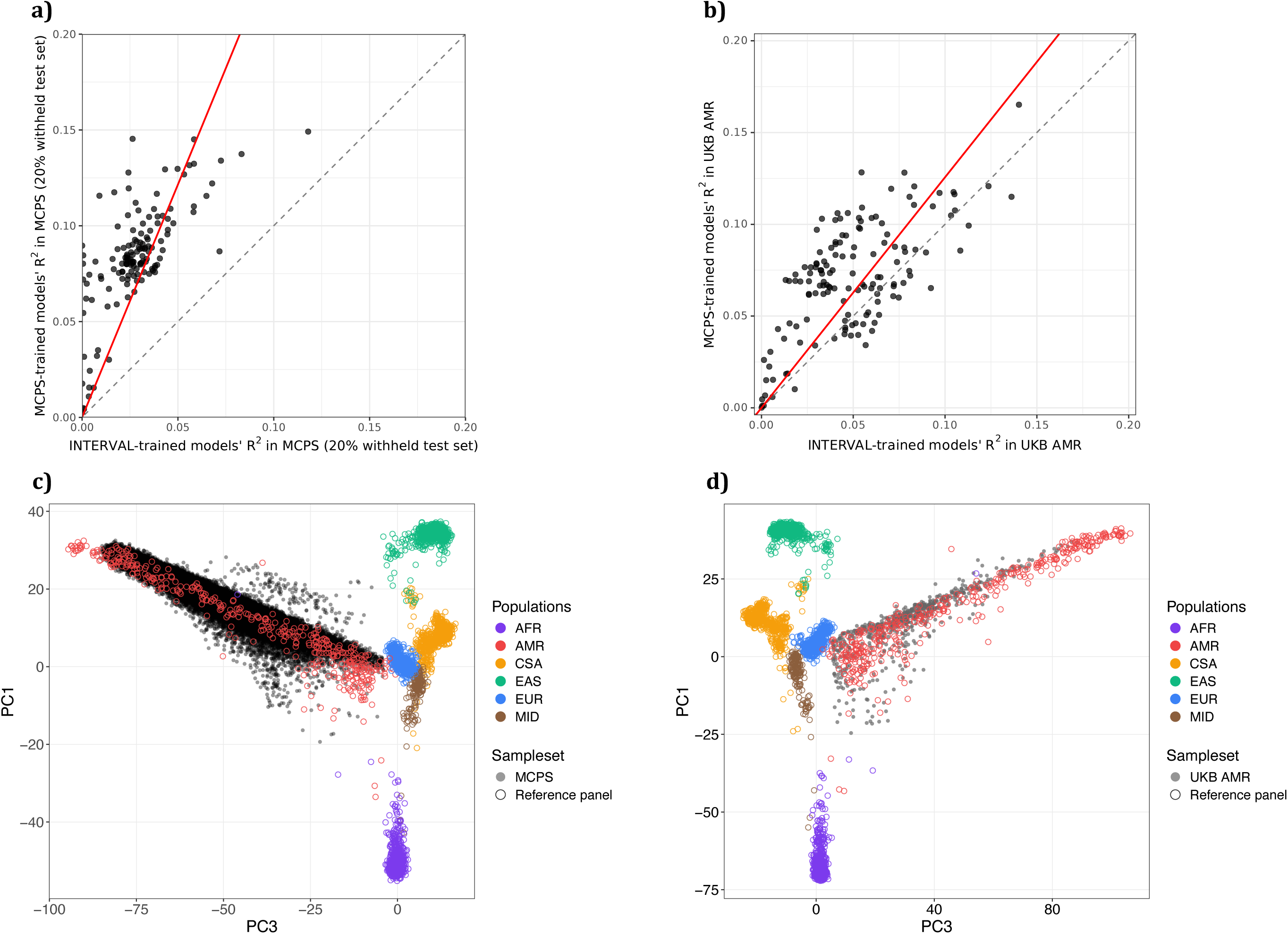
Performance of MCPS- and INTERVAL-trained models in MCPS (a and c) and the AMR subset of the UKB (b and d) Panel **a)** shows the *R*^2^ between each of the 141 measured metabolomic traits and the values predicted by the corresponding Mexico City Prospective Study– (MCPS-) and INTERVAL Study–trained models, as assessed, respectively, in the 20% of the MCPS sample withheld for internal validation and the whole of the MCPS sample (up to *n* = 132,336). The dashed line shows identity in performance; the red line shows the line of best fit through the origin (slope, *λ* = 2.43; 95% confidence interval, CI: 2.26 to 2.60). Panel **b)** shows the performance of the same two sets of models as assessed in the subset of the UK Biobank (UKB) of admixed American (AMR) genetic ancestries (up to *n* = 600); the line of best fit through the origin has slope *λ* = 1.26 (95% CI: 1.17 to 1.34) . Panel **c)** shows participants of the MCPS estimated to be of admixed American (AMR) genetic ancestries with ≥ 90% probability (black-filled dots) projected onto a space defined by the first and the third principal component (PCs) from a principal component analysis of the Human Genome Diversity Project (HGDP) + 1000 Genomes Project (1kGP) reference panel (dots in other colours, as indicated by legend); the principal component analysis was the basis for genetic ancestry classification using a 20-nearest-neighbour algorithm. The continental-level groups in the reference sample are indicated by “AFR” for African, “AMR” for admixed American, “CSA” for Central/South Asian, “EAS” for East Asian, “EUR” for European, and “MID” for Middle Eastern. Panel **d)** shows participants of the UK Biobank (UKB) estimated to be of admixed American (AMR) genetic ancestries with ≥ 90% probability (black-filled dots) projected onto a space defined by the PCs from a principal component analysis of the same reference panel; however, different sets of genetic variants were used for the principal component analyses underlying plots in c) and d), determined by the intersection of the variants in the reference panel with those available in the MCPS and the UKB, respectively, which is why the coordinates of the observations in the reference panel do not align across the two plots.

The predictive performance of MCPS-trained models (**Fig. 2**) among participants in the withheld subset with a higher (≥70%) estimated IAM genetic ancestries proportion (median *R*^2^: 0.082 and IQR: 0.024) was greater than that of INTERVAL-trained models (median *R*^2^: 0.021 and IQR: 0.015) and similar to the MCPS-trained models’ performance in the overall withheld subset. The performance of MCPS-trained models was also improved in the withheld subset with ≥70% estimated EUR genetic ancestries proportion (median *R*^2^: 0.126 and IQR: 0.065 for MCPS-trained models vs. median *R*^2^: 0.054 and IQR: 0.056 for INTERVAL-trained models).

As expected, there was a positive relationship (Pearson’s correlation coefficient, *r* = 0.66) between the metabolomic traits’ estimated heritability (via LD Score regression) and the corresponding models’ predictive performance (**Supplementary Fig. 1**).

### External validation in the UK Biobank and the All of Us Research Program

We validated the new models externally using data from up to 600 UKB participants of AMR genetic ancestries, with metabolomics collected using the same Nightingale Health NMR platform as in MCPS. Models trained in MCPS appeared to be more transferable, with a substantially higher performance in this context (**Fig. 3b** and **Supplementary Table 4**) as compared to that of INTERVAL-trained models (median R^2^ of 0.070 and IQR of 0.039 vs. a median R^2^ of 0.046 and IQR of 0.034, respectively). A direct model comparison revealed MCPS-trained models performed similarly (**Supplementary Fig. 2**) in the AMR-ancestries subset of UKB as in the withheld subset of MCPS (*λ* = 0.85, 95% confidence intervals, CI: 0.81 to 0.88); this slight attenuation may be due to differences in genetic ancestries between the AMR participants of the MCPS and of the UKB (**Fig. 3c–d**), among other differences.

In the subset of AoU participants of AMR genetic ancestries with biomarker and covariate data (between 4,349 and 21,737 participants of age ≥ 35), we again found MCPS-trained models to be more transferable to this context than INTERVAL-trained models (**Fig. 4** and **Supplementary Table 5**). MCPS-trained models had a mean *R*^2^ of 0.035 and standard deviation (s.d.) of 0.032, compared with a mean *R*^2^ of 0.027 and s.d. of 0.023 for INTERVAL-trained models in predicting six AoU traits based on clinical measurements, each matched (**Methods**) to a metabolomic trait measured in MCPS and the UKB using the Nightingale Health NMR spectroscopy platform (the mean rather than the median is presented here due to the small number of estimates considered; *λ* = 1.31; 95% CI: 0.90 to 1.72). However, with the exception of the model for HDL cholesterol, the performance of MCPS-trained models in this subset of AoU, as expected due to differences in the manner in which they were measured (**Methods**), did not match their performance in the withheld subset of MCPS, where, for the equivalent six traits the mean *R*^2^ for MCPS-trained models was 0.065 (s.d.: 0.034) and 0.023 (s.d.: 0.018) for INTERVAL-trained models (**Fig. 4b**).

**Fig. 4:**
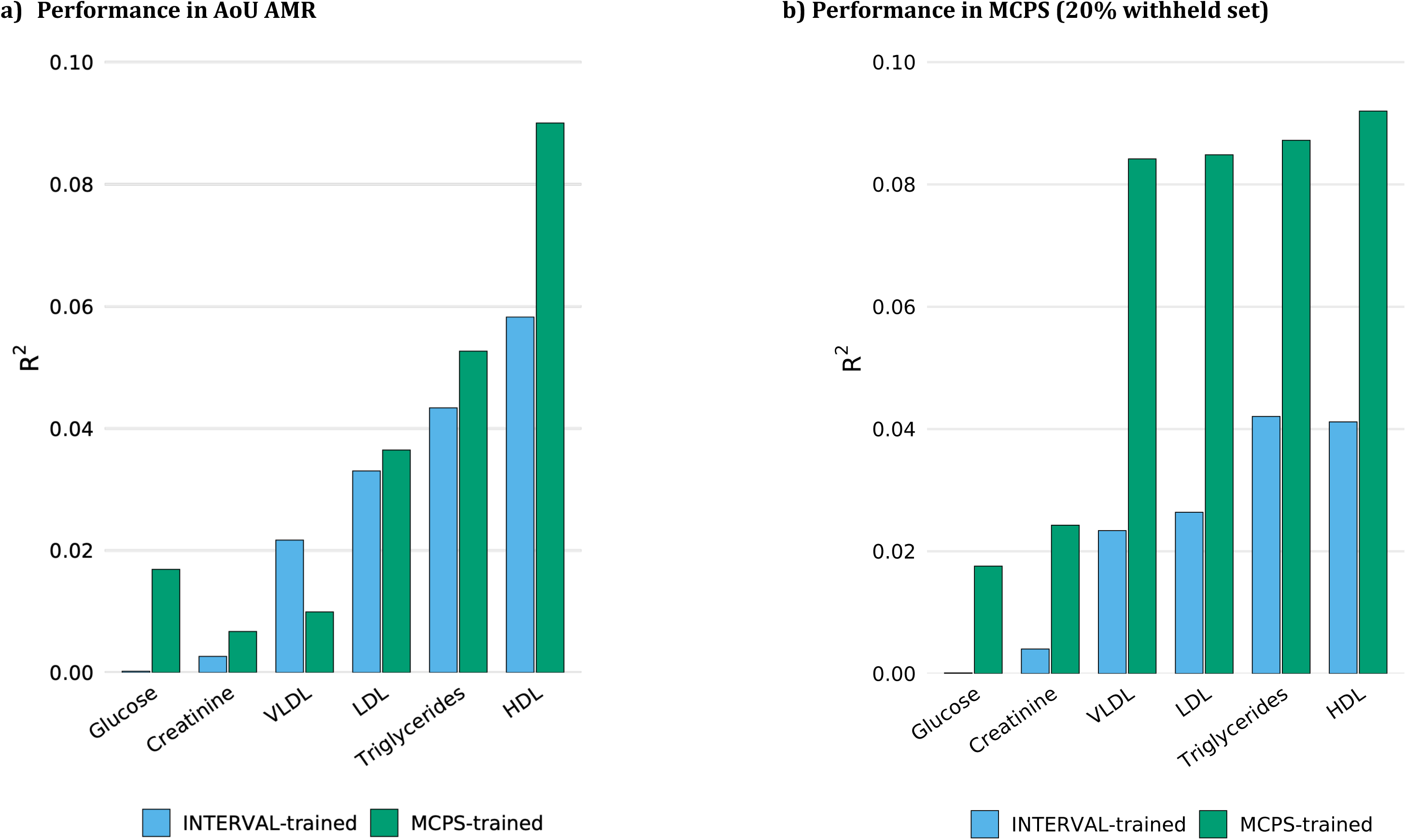
Performance of MCPS- and INTERVAL-trained models for six traits among AoU participants of AMR genetic ancestries and among MCPS participants for the same six traits. **a**, Predictive performance in terms of *R*^2^ of six INTERVAL Study– and Mexico City Prospective Study– (MCPS-)trained models in the subset of the All of Us Research Program (AoU) of admixed American genetic ancestries (AMR), for the six traits available in AoU and matched to traits measured by Nightingale Health’s nuclear magnetic resonance (NMR) spectroscopy platform, which was used in MCPS. **b,** Predictive performance of the same models in MCPS.

### Performance by sex, age group, and health status at baseline across cohorts

Secondary analyses of model performance by sex, age group, and health status at baseline are summarised in **Supplementary Table 8**. Briefly, in MCPS, INTERVAL-trained models had a higher performance among female than among male participants, though this did not explain the differences by ancestry proportion referred to above (**Supplementary Fig. 3**). No notable differences were observed by age group or health status at baseline (**Supplementary Figs. 4– 5**). Similar subgroup analyses for MCPS-trained models in the withheld 20% subset of MCPS again showed higher performance among female than among male participants (**Supplementary Fig. 6**), and higher performance among participants of ages 35–49 and 50–64 than among those of age ≥ 65, with no notable differences by health status at baseline (**Supplementary Figs. 7–8**). Summaries of the models’ performance in subgroups of the external validation samples, the UKB and AoU participants of AMR genetic ancestries, are provided in **Supplementary Table 8**.

### Predicted metabolome–wide association analyses in All of Us

To assess the potential aetiological insights resulting from enhanced metabolome-wide association studies in populations of AMR ancestries, we predicted metabolomic traits in the AMR subset of AoU using both the MCPS- and INTERVAL-trained models, then compared the MWAS associations detected for three high-burden cardiometabolic diseases: type 2 diabetes, ischaemic heart disease, and chronic kidney disease (**Methods; Supplementary Table 6**).

Across the three diseases (**Fig. 5**, **Supplementary Table 9**, and **Supplementary Figs. 9–12**), the MWAS using MCPS-trained models yielded a 5-fold increase in the number of statistically significant associations (FDR-corrected *P* < 0.05) as compared to INTERVAL-trained models: 69 vs. 27 significant associations with type 2 diabetes (with a Pearson’s correlation *r* = 0.58 between the log-odds estimates obtained using the two sets of models), 26 vs. 0 with chronic kidney disease (*r* = 0.65), and 55 vs. 3 with ischaemic heart disease (*r* = 0.71).

**Fig. 5:**
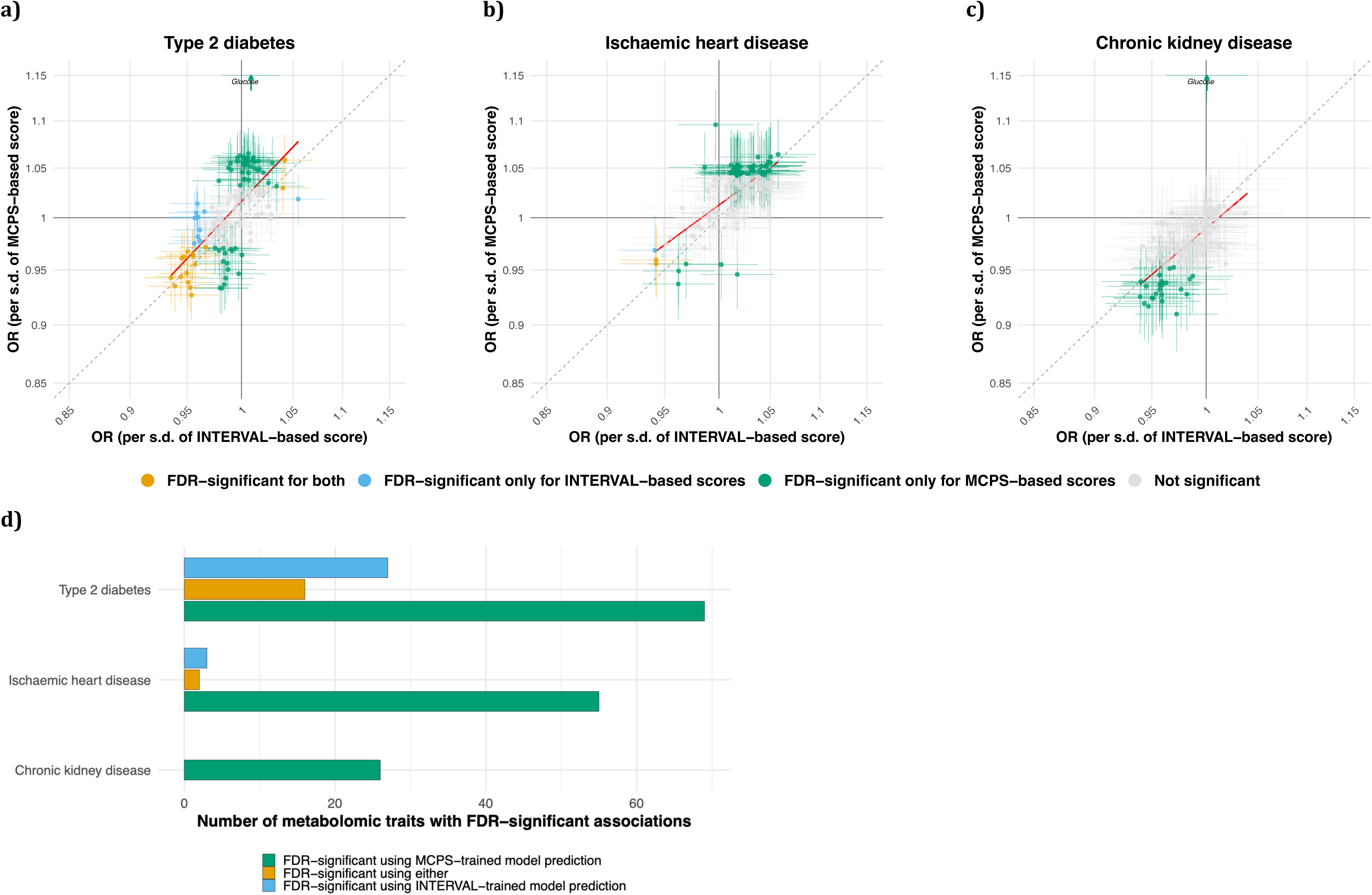
Associations of cardiometabolic diseases with metabolomic traits predicted by MCPS- and INTERVAL-trained models. The estimated associations of 139 metabolomic traits, predicted using two different sets of models, with three high-burden cardiometabolic diseases (log-odds and 95% confidence intervals plotted with axis tick labels exponentiated to represent odds ratios, ORs, per standard deviation difference in the genetically predicted trait): **a)** type 2 diabetes; **b)** ischaemic heart disease; **c)** chronic kidney disease. The estimates obtained using traits predicted by the MCPS-trained models (y-axis) are plotted against the estimates obtained using traits predicted by the INTERVAL-trained models (x-axis). Associations were estimated using logistic regression adjusting for age at the last electronic health record event, sex, and the first 10 principal genetic components. Point colours indicate statistical significance after applying false discovery rate (FDR) correction for multiple testing within each disease and model set (MCPS-vs. INTERVAL-trained): significant using both sets of models (orange), significant using only MCPS-based prediction (green), significant using only INTERVAL-based prediction (blue), or not significant (grey). The dashed line represents the identity line. The red line represents the linear regression fit of the MCPS vs. INTERVAL log-odds estimates. In panels **a)** and **c)**, the strong association of glucose predicted by the MCPS-trained model with, respectively, type 2 diabetes and chronic kidney disease, extends beyond the upper limit of the y-axis and is indicated by an upward-pointing arrow. Panel **d)** summarises the overall number of FDR-significant associations estimated per disease, using each set of models (and the overlap, i.e. the number of FDR-significant associations estimated using both sets of models).

The analyses detected multiple known biomarker–disease associations. For example, circulating glucose, as predicted for AoU participants of AMR genetic ancestries by the MCPS-trained model, was strongly associated with type 2 diabetes (OR: 1.34 per standard deviation, s.d.; 95% CI: 1.31 to 1.38; FDR-corrected *P* = 8.45 × 10^−107^), as expected given the role of elevated glucose in the definition of the disease. Notably, however, glucose as predicted by the INTERVAL-trained model was not associated with type 2 diabetes (OR: 1.01 per s.d.; 95% CI: 0.98 to 1.03; FDR-corrected *P* = 0.76). Glucose as predicted by the MCPS-trained model was also statistically significantly associated with ischaemic heart disease (OR: 1.10; 95% CI: 1.06 to 1.13; FDR-corrected *P* = 2.88 × 10^−5^) and chronic kidney disease (OR: 1.16; 95% CI: 1.12 to 1.21; FDR-corrected *P* = 1.98 × 10^−13^), whereas analyses of glucose predicted by the INTERVAL-trained model yielded no significant associations with the two diseases (FDR-corrected *P* ≥ 0.76; ORs 0.99–1.00)^54–56^.

Total triglycerides predicted by MCPS-trained models was likewise statistically significantly associated with type 2 diabetes (OR: 1.06; 95% CI: 1.03 to 1.09; FDR-corrected *P* = 4.15 × 10^−5^) and ischaemic heart disease (OR: 1.05; 95% CI: 1.01 to 1.08; FDR-corrected *P* = 0.036), while total triglycerides predicted by INTERVAL-trained models was not (FDR-corrected *P* ≥ 0.56; ORs both 1.01)^57–59^.

The MWAS conducted using MCPS-trained models in the AMR subset of AoU also corroborated biomarker–disease associations detected in previous analyses of NMR metabolomic data in both the UKB and MCPS^60–62^. For example, triglyceride concentrations in 15 of the 17 lipoprotein classes and subclasses predicted by MCPS-trained models in AoU were statistically significantly associated with type 2 diabetes, whereas only two of the 17 equivalent traits predicted by INTERVAL-trained models had significant associations with type 2 diabetes (**Supplementary Table 9** and **Supplementary Fig. 10**). Similarly, free cholesterol in large, very large, and chylomicrons and extremely large VLDL (predicted using MCPS-trained models) were positively and statistically significantly associated with type 2 diabetes (all ORs 1.06; FDR-corrected *P* ≤ 1.44 × 10^−4^). Particle concentrations of large VLDL, very large VLDL, and extremely large VLDL as predicted by MCPS-trained models were also associated with type 2 diabetes (ORs 1.06–1.07; FDR-corrected *P* ≤ 1.07 × 10^−4^), as were phospholipids in the same VLDL subclasses (all ORs 1.06; FDR-corrected *P* ≤ 1.44 × 10^−4^).

Furthermore, several associations between predicted lipoprotein subclass traits and ischaemic heart disease in the AMR subset of AoU (when using MCPS-trained models to predict the traits) were consistent with previously reported UKB estimates^61^ and estimates for vascular occlusive mortality in MCPS, which predominantly comprised mortality from ischaemic heart disease)^60^. These included (**Supplementary Fig. 11**) concentrations of medium, large, very large, and extremely large VLDL (ORs 1.04–1.05; FDR-corrected *P* ≤ 0.043), as well as phospholipids in small, medium, large, very large, and extremely large VLDL (ORs 1.04–1.05; FDR-corrected *P* ≤ 0.040). Associations with triglycerides in IDL and in large, medium, and small LDL (ORs 1.04– 1.05; FDR-corrected *P* ≤ 0.040) were also consistent with UKB estimates, although the corresponding previously reported estimates for vascular-occlusive mortality in MCPS were strongest for triglycerides in large LDL and not statistically significant for the other subclasses.

Consistently with previously reported analyses of kidney function and NMR metabolomic traits in MCPS^63^, several HDL-related traits in the AMR subset of AoU (predicted using MCPS-trained models) were inversely associated with chronic kidney disease (**Supplementary Fig. 12**). These included small HDL particle concentration, free cholesterol in small HDL, phospholipids in small HDL, and total lipids in small HDL (ORs 0.91–0.94; FDR-corrected *P* ≤ 0.012).

Finally, not all associations of metabolomic traits predicted by MCPS-trained models were directionally concordant with the previously reported estimates. Free cholesterol and phospholipids in small HDL, for instance, were inversely associated with type 2 diabetes (ORs 0.95–0.96 and FDR-corrected *P* ≤ 0.002) in the AMR-ancestries subset of AoU, in contrast with previously reported positive estimates based on UKB and MCPS data^60,61^.

## Discussion

In this work, we sought to enable metabolome-wide association studies in cohorts of admixed American ancestries, thereby helping to reduce inequities in multi-omics research in Latin America and populations of AMR ancestries more broadly. We developed genetic scores for metabolomic traits which addressed the performance gap between groups of European and admixed American ancestries, then performed MWAS showing a multiple-fold increase in power to detect metabolite-disease associations. Our MWAS detected metabolic aetiologies in ischaemic heart disease, chronic kidney disease, and type 2 diabetes—diseases which disproportionately burden populations of admixed American ancestries. To empower metabolomics research for AMR populations, we [will] have made our AMR ancestries–trained scores openly available at the OmicsPred^29^ portal (www.omicspred.org).

Metabolomic traits are often key mediators in cardiometabolic disease processes, and exclusive reliance on samples of EUR ancestries risks systematically failing to recognise mechanisms that disproportionately manifest in non-EUR populations as well as, consequently, potential treatments^1,3,7,36^. For AMR populations, this is particularly important in metabolism research, since previous work has highlighted the heavy burden and high prevalence of undertreated metabolic disease^11–13^. The discovery of genetic variants strongly associated with type 2 diabetes in populations with AMR genetic ancestries but are rare or absent elsewhere also make clear the need for ancestry-specific analyses^14–16^. Developing genetic prediction models for metabolomic traits tailored to populations of diverse genetic ancestries is therefore a step toward research equity and the equity of downstream health benefits from multi-omics research^8,64^.

We showed that current genetic scores trained on European ancestries for metabolome-wide association studies have poor transferability to admixed American ancestries. This attenuation in performance in AMR ancestries was not only evident relative to the performance observed in samples of EUR ancestries but also more pronounced than that in other non-EUR samples^25^. In contrast, training models for metabolomic traits within a large cohort of AMR genetic ancestries, the Mexico City Prospective Study^28,30^, yielded substantial improvements in internal validation performance, consistent with previous research on the sensitivity of genetic scores to linkage disequilibrium and allele frequency differences across genetic ancestry groups^27,65^. These findings were supported by external validation in AMR-ancestries subsets of the UK Biobank and the All of Us Research Program.

These gains in predictive performance translated into greater power for biomarker discovery for three cardiometabolic diseases with high burden in AMR populations. In the large AMR subset of AoU, the MWAS based on MCPS-trained models identified the inherent relationship between glucose and type 2 diabetes and replicated the association between total triglycerides with ischaemic heart disease, neither of which were detected when using models trained on EUR-ancestries samples^54–59^. Beyond these established relationships, the MWAS based on MCPS-trained models also provided additional evidence for associations detected in previous analyses of measured NMR metabolomic traits in UKB and MCPS^60–63^, including associations of type 2 diabetes with finer triglyceride, free cholesterol, particle concentration, and phospholipid measures across several lipoprotein subclasses, and associations of chronic kidney disease with lower HDL-related measures. For ischaemic heart disease, VLDL subclass particle and phospholipid associations detected in the AMR subset of AoU were also consistent with previously reported UKB estimates as well as MCPS analyses of vascular occlusive mortality. Overall, these results show that ancestry-matched prediction can extend fine-scale metabolomic epidemiology to AMR populations.

In addition to providing a new set of tools for metabolomic research in AMR populations, this work provides a strong rationale for metabolomic profiling in subsets of cohorts with participants of other underrepresented genetic ancestries. Such an approach may allow for more effective prediction of omic traits for the remainder of the cohort and, as shown here for the AMR subset of AoU, for ancestry-specific subsets of other diverse cohorts. Equity in downstream analyses is contingent upon improved equity in the training data.

Our study has several limitations. First, while the models exhibited improved performance across three diverse AMR samples, AMR is a broad category, and, as such, does not take into account finer genetic structure that varies within and between AMR populations in Mexico (where the training data in this study were generated), the Americas, and elsewhere^28,64^. Extending the argument from this work, we would expect performance of the new genetic models for metabolomic trait prediction to vary across populations of AMR genetic ancestries with different histories and in different environments, as is already shown here within the two cohorts used in external validation of the models. Second, as genetic prediction models only capture the heritable component of the metabolome, results of downstream analyses, by design, do not reflect other, environmental factors that influence both metabolic biomarker levels and disease risk.

In addition to making use of the newly released genetic prediction models for metabolomic traits in association analyses and other metabolomic research, future work could explore the development of AMR-specific models for other omic layers, such as the proteome and transcriptome, subject to the availability of training data. Incorporating individual-level information on local genetic ancestry might provide further improvements in the performance of genetic prediction models of metabolomic traits^66,67^.

In summary, we have shown that existing genetic scores for metabolomic traits developed in a sample of predominantly European genetic ancestries fall short in populations of admixed American ancestries. The training of new models in a cohort of AMR ancestries provides substantial gains in predictive performance and, crucially, in the power of downstream association analyses to elucidate metabolic aetiology. These findings strongly indicate that the expansion of metabolomic profiling to diverse individuals of underrepresented populations is necessary for equitable understanding of metabolic disease.

## Data availability

This study used data from the Mexico City Prospective Study Resource, from the UK Biobank Resource (under Application Number 31461), and the *All of Us* Research Program’s Controlled Tier Dataset version crdv8 – R9 (through the workspace with ID aou-rw-f84dbec1). Data from the Mexico City Prospective Study are available to *bona fide* academic researchers. For more details, the study’s Data and Sample Sharing policy may be viewed (in English or Spanish) at https://www.ctsu.ox.ac.uk/research/mcps. Available study data can be examined in detail through the study’s Data Showcase, available at https://datashare.ndph.ox.ac.uk/mexico/. MCPS ancestry-specific allele frequencies are available in a public browser (https://rgc-mcps.regeneron.com/). UK Biobank data are available for *bona fide* researchers to conduct health-related research that is in the public interest through the UK Biobank Research Analysis Platform. Available study data can be examined in detail through the study’s Data Showcase, available at https://biobank.ndph.ox.ac.uk/showcase/. All of Us Research Program data is available to authorised users on the Researcher Workbench (https://workbench.researchallofus.org/login).

The MCPS-trained models (genetic scores) developed in this study are publicly accessible through the OmicsPred portal (https://www.omicspred.org/) under accession codes OPGS3339470–OPGS3339610. The INTERVAL Study–trained genetic models used in this study used to develop genetic scores are publicly accessible through the OmicsPred portal (https://www.omicspred.org/) under accession codes OPGS003419–OPGS003559.

## Code availability

The original code used to train the genetic models for metabolomic traits with MCPS data, internally validate them and compare them with INTERVAL-trained models, externally validate them using UK Biobank and All of Us Research Program data, and perform association analyses of predicted metabolomic traits with diseases, is available at https://github.com/tinoreskovic/omicspred_amr_metabolomics.

## Acknowledgments

The Mexico City Prospective study (MCPS) is a long-standing scientific collaboration between researchers at the National Autonomous University of Mexico and the University of Oxford and has received funding from the Mexican Health Ministry; the National Council of Science and Technology for Mexico; Wellcome [058299/Z/99]; Cancer Research UK; the British Heart Foundation [RE/13/1/30181]; Kidney Research UK [MR/R007764/1]; and the UK Medical Research Council [MC_UU_00017/2, MR/Z504543/1]. Genotyping in MCPS was funded through an academic partnership between the National Autonomous University of Mexico, the University of Oxford, Regeneron and AstraZeneca. This work was conducted using the MCPS Resource, as well as the UK Biobank Resource (under Application Number 31461), and the *All of Us* Research Program’s Controlled Tier Dataset version crdv8 – R9 (through the workspace with ID aou-rw-f84dbec1). We gratefully acknowledge MCPS, UK Biobank, and *All of Us* participants for their contributions, without whom this research would not have been possible. We also thank the National Institutes of Health’s *All of Us Research Program* for making available the participant data examined in this study. T.O. is supported by a Health Data Research UK Early Career Research Fellowship, specifically as part of the HDR UK Molecules to Health Records Driver Programme (HDRUK.2023.0028). Health Data Research UK is funded by UK Research and Innovation, the Medical Research Council, the British Heart Foundation, Cancer Research UK, the National Institute for Health and Care Research, the Economic and Social Research Council, the Engineering and Physical Sciences Research Council, Health and Care Research Wales, Health and Social Care Research and Development Division (Public Health Agency, Northern Ireland), Chief Scientist Office of the Scottish Government Health and Social Care Directorates. These funding sources had no role in the design, conduct or analysis of the study or the decision to submit the manuscript for publication. C.F., X.K., Y.X. & MI. were supported by core funding from the British Heart Foundation (RG/F/23/110103), NIHR Cambridge Biomedical Research Centre (NIHR203312) [*], BHF Chair Award (CH/12/2/29428) and the Cambridge BHF Centre of Research Excellence (RE/24/130011). M.I. is also supported by the Munz Chair of Cardiovascular Prediction and as well as by the UK Economic and Social Research 878 Council (ES/T013192/1). Participants in the INTERVAL randomised controlled trial were recruited with the active collaboration of NHS Blood and Transplant England (www.nhsbt.nhs.uk), which has supported field work and other elements of the trial. DNA extraction and genotyping were co-funded by the National Institute for Health and Care Research (NIHR), the NIHR BioResource (http://bioresource.nihr.ac.uk) and the NIHR Cambridge Biomedical Research Centre (BRC-1215-20014) [*]. The academic coordinating centre for INTERVAL was supported by core funding from the: NIHR Blood and Transplant Research Unit (BTRU) in Donor Health and Genomics (NIHR BTRU-2014-10024), NIHR BTRU in Donor Health and Behaviour (NIHR203337), UK Medical Research Council (MR/L003120/1), British Heart Foundation (SP/09/002; RG/13/13/30194; RG/18/13/33946), NIHR Cambridge BRC (BRC-1215-20014; NIHR203312) [*], and by Health Data Research UK, which is funded by the UK Medical Research Council, Engineering and Physical Sciences Research Council, Economic and Social Research Council, Department of Health and Social Care (England), Chief Scientist Office of the Scottish Government Health and Social Care Directorates, Health and Social Care Research and Development Division (Welsh Government), Public Health Agency (Northern Ireland), British Heart Foundation and Wellcome. A complete list of the investigators and contributors to the INTERVAL trial is provided in reference [**]^38^. The academic coordinating centre would like to thank blood donor centre staff and blood donors for participating in the INTERVAL trial.

*The views expressed are those of the authors and not necessarily those of the NIHR or the Department of Health and Social Care.

**Di Angelantonio E, Thompson SG, Kaptoge SK, Moore C, Walker M, Armitage J, Ouwehand WH, Roberts DJ, Danesh J, INTERVAL Trial Group. Efficiency and safety of varying the frequency of whole blood donation (INTERVAL): a randomised trial of 45 000 donors. Lancet. 2017 Nov 25;390(10110):2360-2371.

## Author information

### Contributions

T.O., D.J., J.M.T. and M.I. conceived and designed the study. T.O. performed the training, internal validation, and external validation of MCPS-trained models, comparisons with the INTERVAL-trained models, and downstream association analyses with diseases. T.O. and D.J. performed the MCPS validation of the INTERVAL-trained models. E.T. and D.A.R. performed the GWAS of metabolomic traits in MCPS. S.L., J.R.E., J.M.T., and M.I. jointly supervised this work. T.O. wrote the original manuscript draft. All authors contributed to the work, including through data generation and curation, analytical support, interpretation of findings, and revision of the manuscript. All authors reviewed and approved the final manuscript.

### Ethics declarations

M.I. is a trustee of the Public Health Genomics (PHG) Foundation and a member of the Scientific Advisory Boards of Open Targets and CIC bioGUNE. He has research collaborations with AstraZeneca and Nightingale Health Ltd. J.R.E declares grants to the University of Oxford from Regeneron and AstraZeneca. S.L. reports receiving grants from the UK Medical Research Council, the US Centers for Disease Control and Prevention Foundation (with financial support from Amgen) and the World Health Organization, HDR UK (grant no. HDRUK2023.0028) funded by the MRC, Engineering and Physical Sciences Research Council, Economic and Social Research Council, Department of Health and Social Care (England), Chief Scientist Office of the Scottish Government Health and Social Care Directorates, Health and Social Care Research and Development Division (Welsh Government), Public Health Agency (Northern Ireland), BHF and Cancer Research UK. The Clinical Trial Service Unit and the Epidemiological Studies Unit at the University of Oxford receives research grants from industry that are governed by University of Oxford contracts, which protects the independence of the Clinical Trial Service Unit and the Epidemiological Studies Unit. D.J. is currently employed by Boehringer Ingelheim. This employment is unrelated to the work presented in this manuscript, which was conducted while D.J. was affiliated with the University of Oxford, the University of Cambridge, and HDR UK; D.J. declares no competing interests related to this work.

### Rights Retention Statement

For the purpose of Open Access, the authors have applied a CC BY public copyright licence to any Author Accepted Manuscript (AAM) version arising from this submission.

## Extended data figures and tables

**Extended Data Fig. 1:**
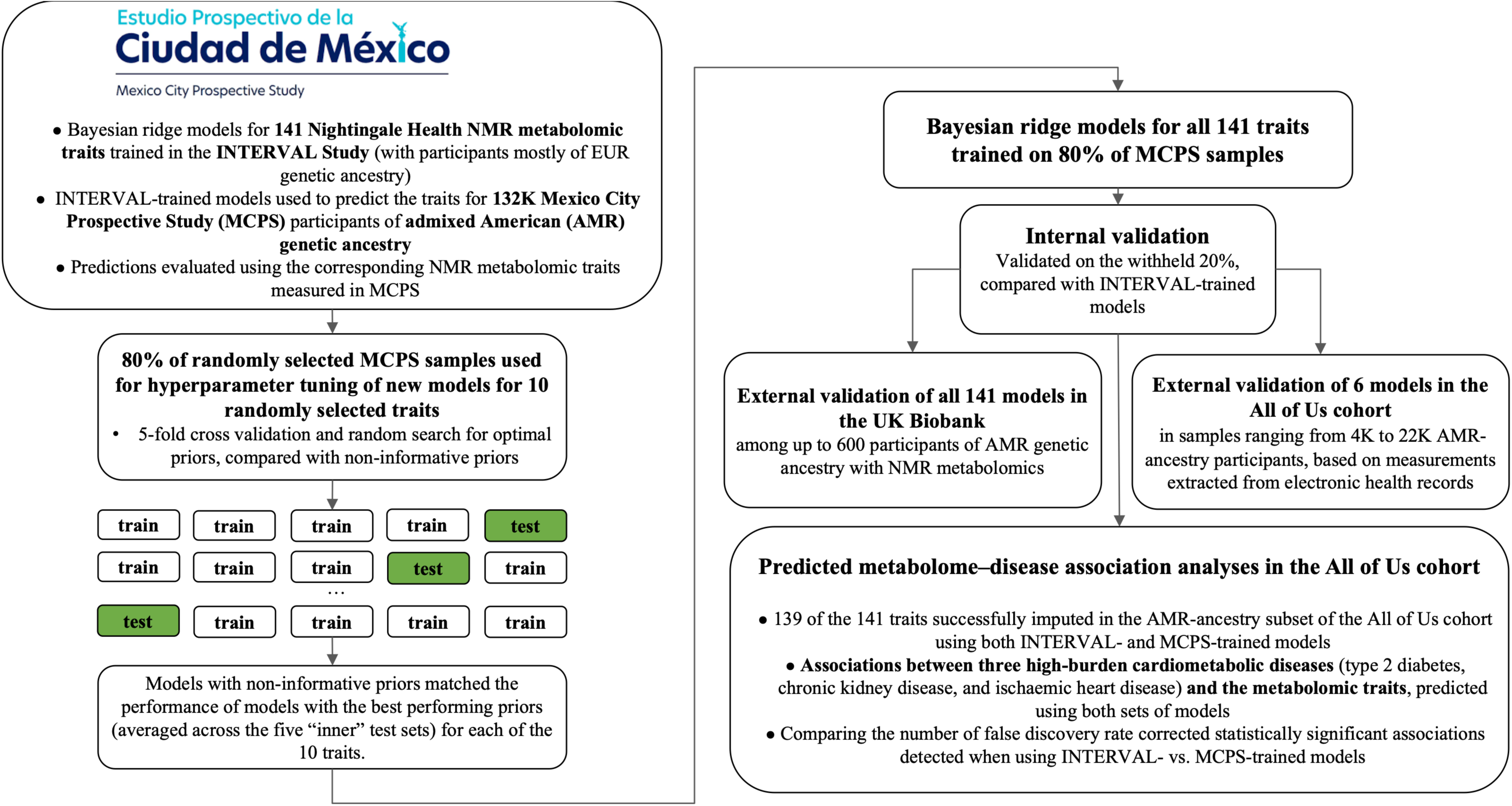
Schematic framework for the validation of INTERVAL Study–trained genetic models for metabolomic traits and the development and validation of new, MCPS-trained genetic models for metabolomic traits in the Mexico City Prospective Study, the UK Biobank, and the All of Us Research Program. This figure shows the overall study design for: (i) the evaluation of INTERVAL Study–trained genetic models for the prediction of metabolomic traits measured via the Nightingale Health nuclear magnetic resonance (NMR) spectroscopy platform in the Mexico City Prospective Study (MCPS); (ii) the development and evaluation of new genetic models for the same traits, trained in MCPS as a large prospective cohort of participants of admixed American (AMR) genetic ancestries with genetic data and metabolomic data generated using the same Nightingale platform; (iii) the external validation of the new MCPS-trained models and comparison of performance with that of INTERVAL-trained models in AMR subsets of the UK Biobank (UKB) and the All of Us Research Program (AoU); and (iv) downstream association analyses of three cardiometabolic diseases with the metabolomic traits predicted using the two sets of models, also in an AMR subset of AoU.

## Supplementary information

Supplementary Figures (download PDF)

Supplementary Figs. 1–12.

Reporting Summary (download PDF)

Supplementary Tables (download xlsx)

Supplementary Tables. 1–9.

