## Supplementary figures for "Enhanced power and transferability for genetics-driven metabolomic biomarker discovery in admixed American cohorts"

#### Supplementary Fig. 1: Relationship between estimated heritability and predictive performance of MCPS-trained models

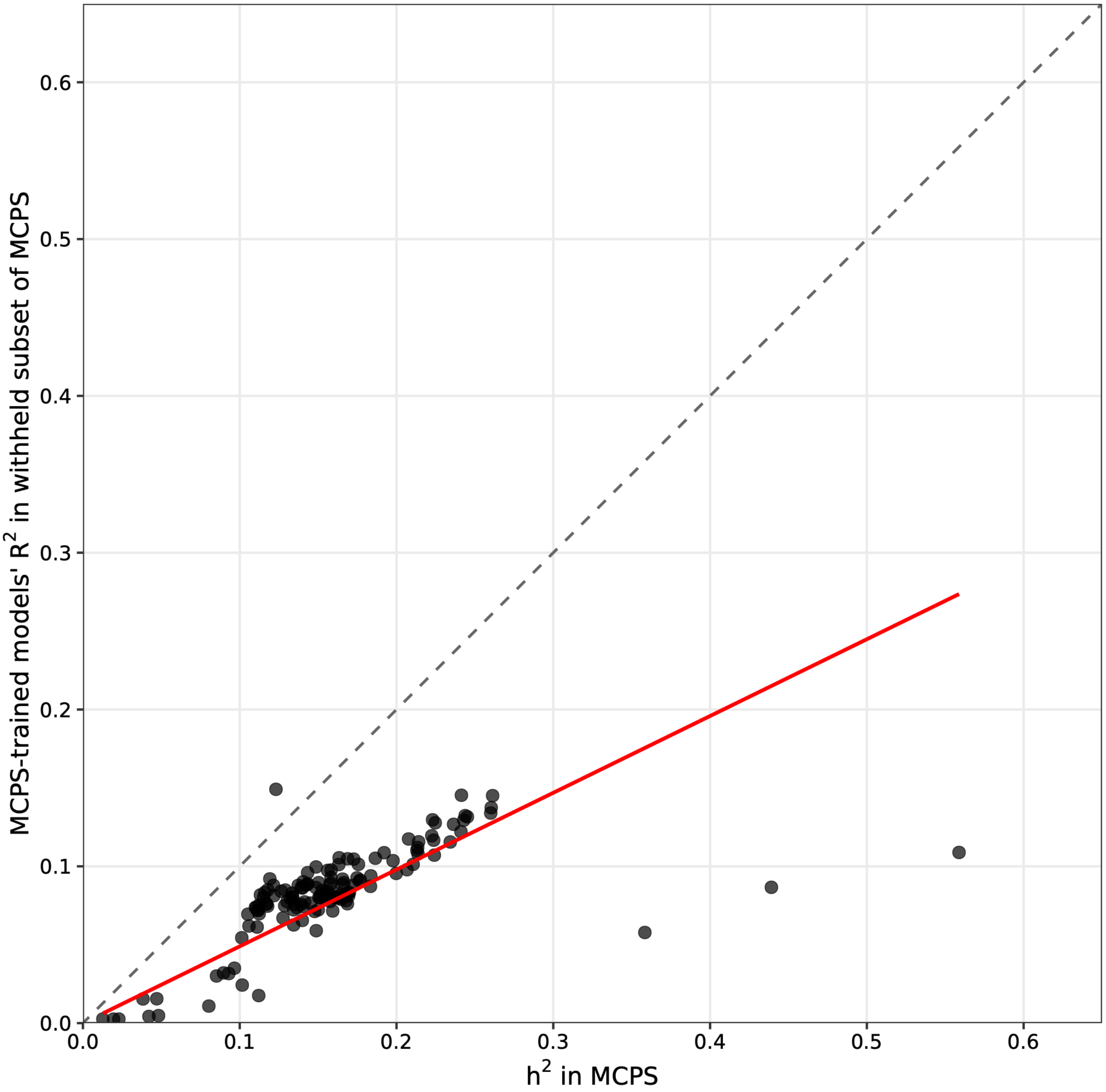


The scatter plot shows the relationship between the estimated heritability of metabolomic traits and the predictive performance of the corresponding Mexico City Prospective Study (MCPS)-trained models. Predictive performance was positively correlated with estimated heritability (Pearson's correlation coefficient r = 0.66); the slope of the line of best fit through the origin was 0.49.

**Supplementary Fig. 2: Predictive performance of MCPS-trained models in the withheld MCPS subset and in the AMR-ancestries subset of the UKB**

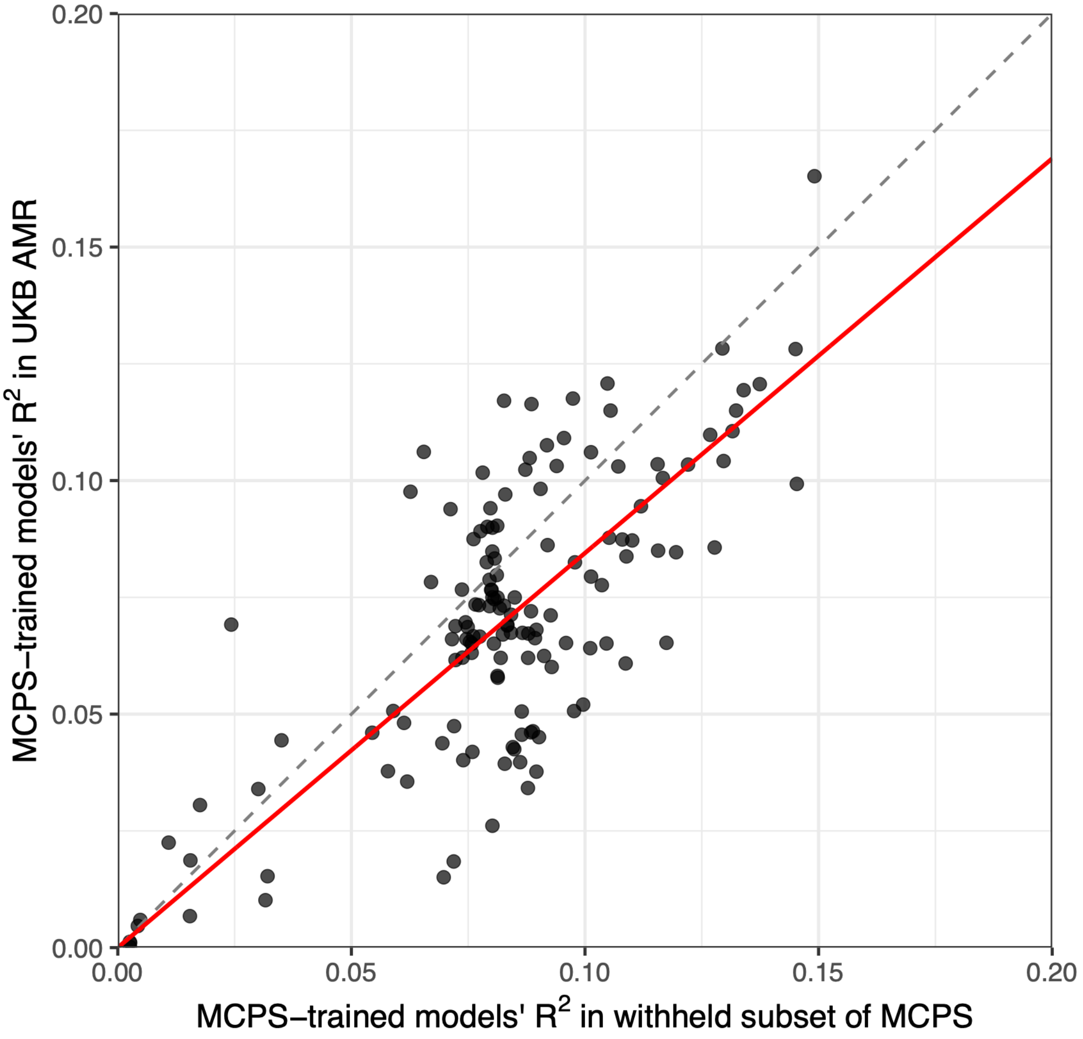


The scatter plot compares the predictive performance, in terms of $R^2$, of Mexico City Prospective Study (MCPS)-trained models evaluated in the 20% of MCPS participants withheld for internal validation and in the subset of the UK Biobank (UKB) of admixed American (AMR) genetic ancestries. The dashed line shows identity in performance; the red line shows the line of best fit through the origin (slope, $\lambda$ = 0.85; 95% confidence interval: 0.81 to 0.88). Median $R^2$ was 0.083 (IQR: 0.021) in the withheld MCPS subset and 0.070 (IQR: 0.039) in UKB AMR.

**Supplementary Fig. 3: Performance of INTERVAL-trained models in MCPS by sex and genetic ancestries proportion**

**a)**

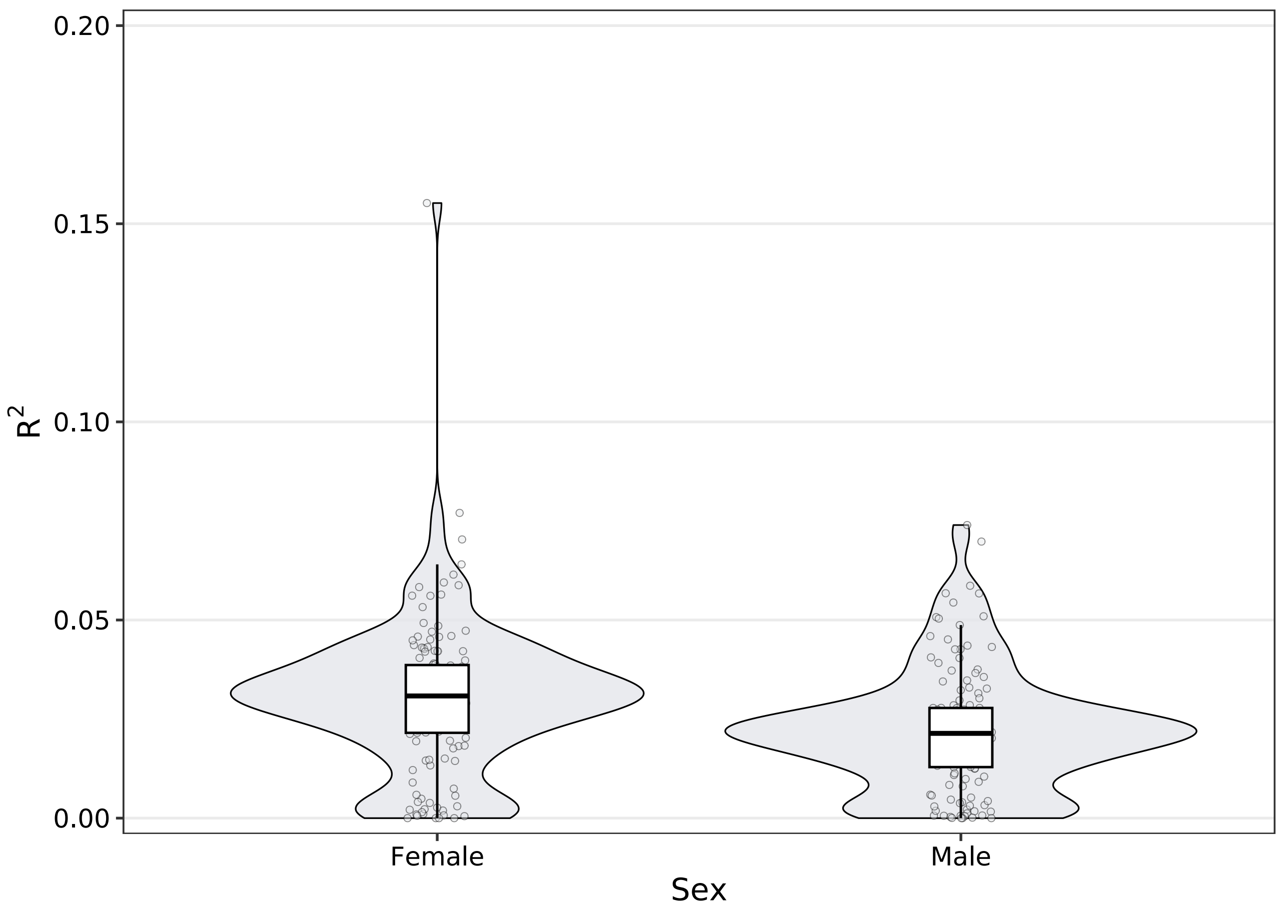


**b)**

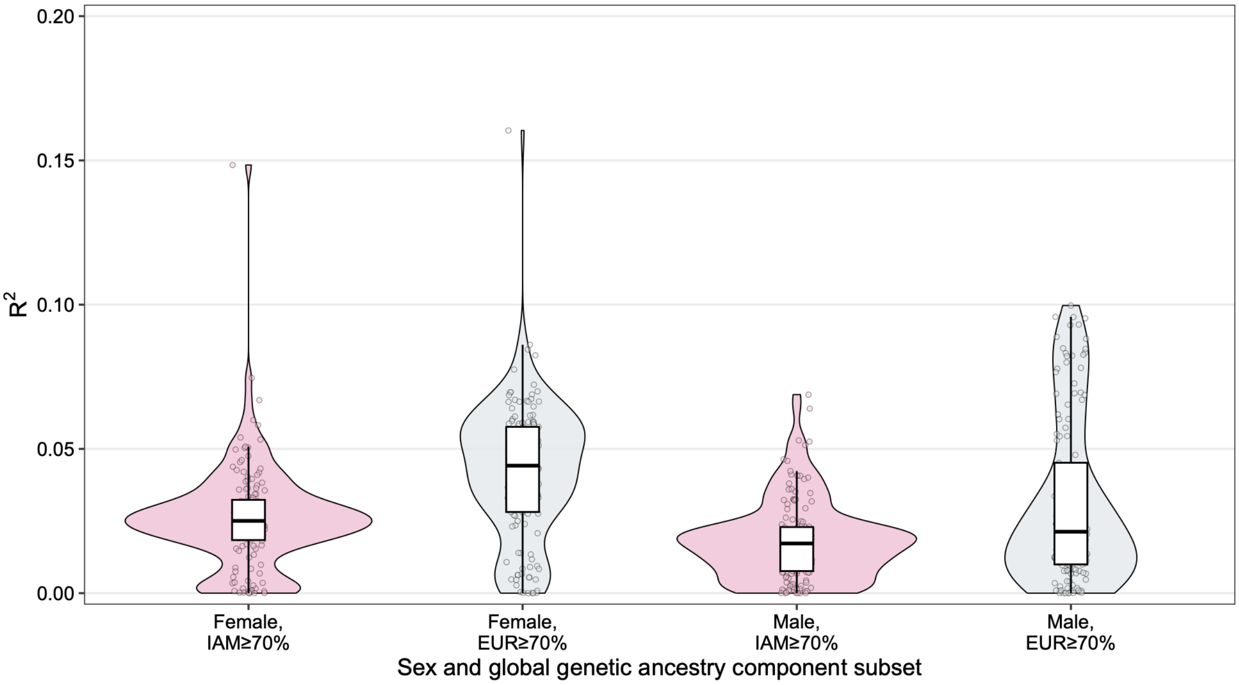


The violin plots and overlaid boxplots show the distribution of $R^2$ values for 141 metabolomic traits predicted by INTERVAL Study–trained models in the Mexico City Prospective Study (MCPS), stratified by sex **a)** and sex as well as **b)** genetic ancestries proportion, i.e., further subsets of the male or female participants of the MCPS defined by higher Indigenous American (IAM) ancestries proportion (≥ 70%) and higher European (EUR) genetic ancestries proportion (≥ 70%). The boxplots show the median, interquartile range, 1.5 times the interquartile range, and outliers; points show the values for individual metabolomic traits. Performance was better among female participants (median $R^2$: 0.031; IQR: 0.017) than among male participants (median $R^2$ 0.021; IQR: 0.015). In panel b, median $R^2$ was 0.025 (IQR: 0.014) among female participants with ≥70% IAM ancestries proportion and 0.044 (IQR: 0.030) among those with ≥70% EUR ancestries proportion; corresponding values among male participants were 0.017 (IQR: 0.015) and 0.021 (IQR: 0.035), respectively.

**Supplementary Fig. 4: Predictive performance of INTERVAL-trained models in MCPS by age group**

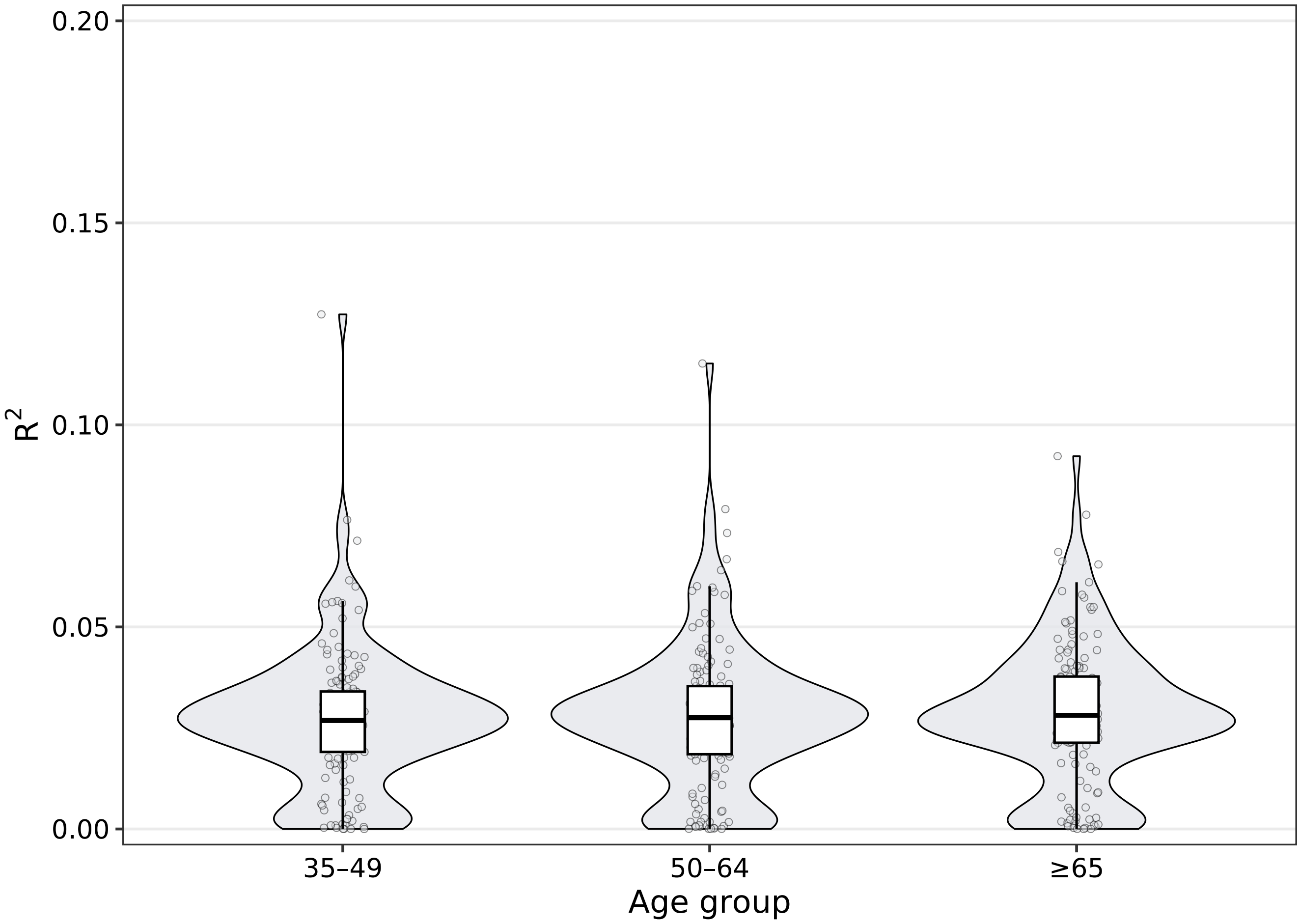


The violin plots and overlaid boxplots show the distribution of $R^2$ values for 141 metabolomic traits predicted by INTERVAL Study-trained models in the Mexico City Prospective Study (MCPS), stratified by age at baseline. The boxplots show the median, interquartile range, 1.5 times the interquartile range, and outliers; points show the values for individual metabolomic traits. Predictive performance was similar across age groups: median $R^2$ was 0.027 (IQR: 0.015) among participants aged 35–49 years, 0.028 (IQR: 0.017) among participants aged 50–64 years, and 0.028 (IQR: 0.016) among participants aged ≥65 years.

**Supplementary Fig. 5: Predictive performance of INTERVAL-trained models in MCPS by diabetes and baseline health status**

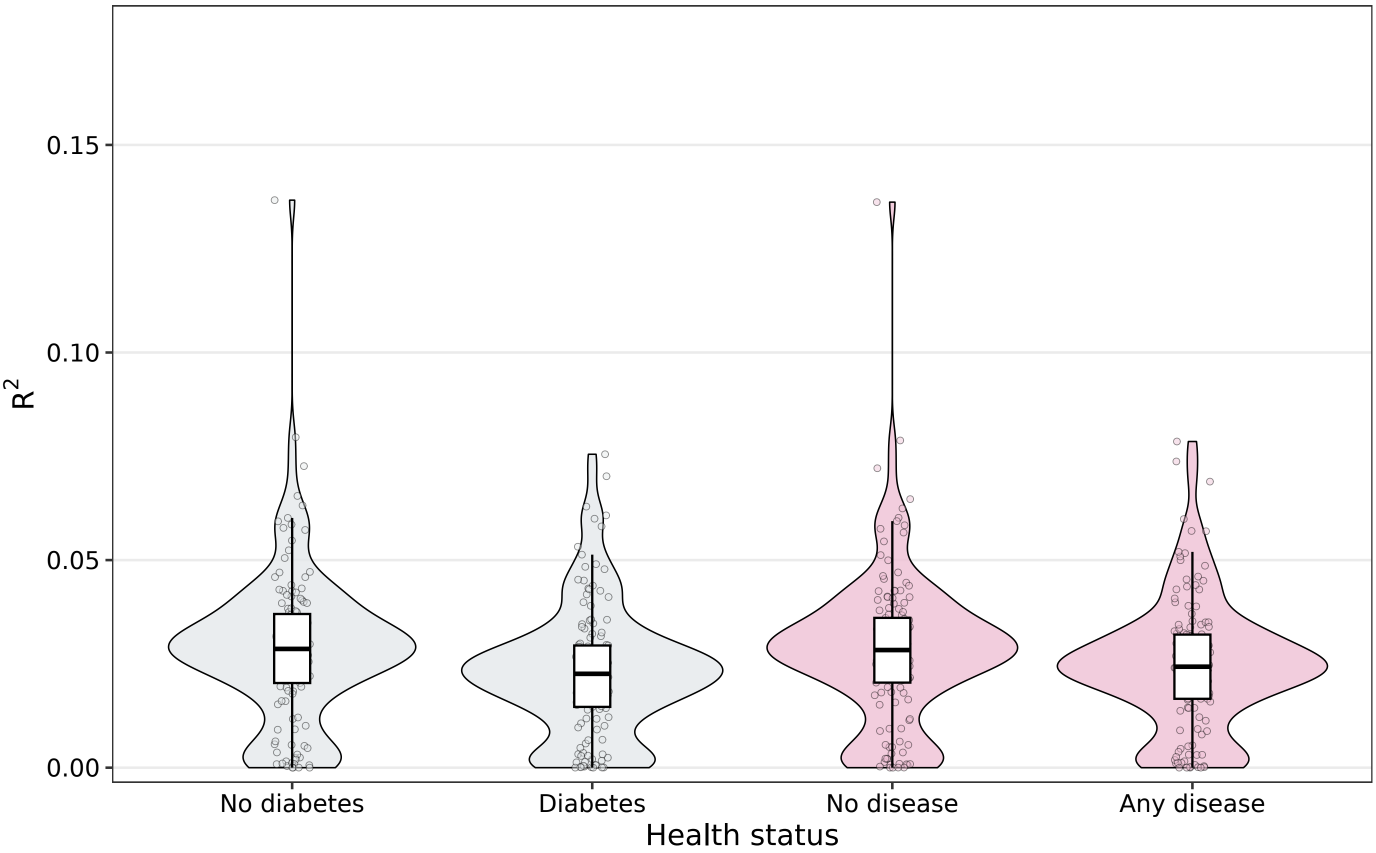


The violin plots and overlaid boxplots show the distribution of $R^2$ values for 141 metabolomic traits predicted by INTERVAL Study-trained models in the Mexico City Prospective Study (MCPS), stratified by diabetes status and by the presence or absence of any of a range of common diseases at baseline. Diabetes was defined as self-reported baseline diabetes or HbA1c >6.5%. No disease denotes participants without baseline cardiovascular disease, cancer, chronic kidney disease, emphysema, cirrhosis, peptic ulcer disease, peripheral arterial disease, diabetes, or HbA1c >6.5%; any disease denotes the complement. Median $R^2$ was 0.029 (IQR: 0.017) among participants without diabetes and 0.023 (IQR: 0.015) among participants with diabetes; it was 0.028 (IQR: 0.016) among participants with no disease and 0.024 (IQR: 0.015) among participants with at least one disease.

**Supplementary Fig. 6: Predictive performance of MCPS-trained models in the withheld MCPS subset by sex**

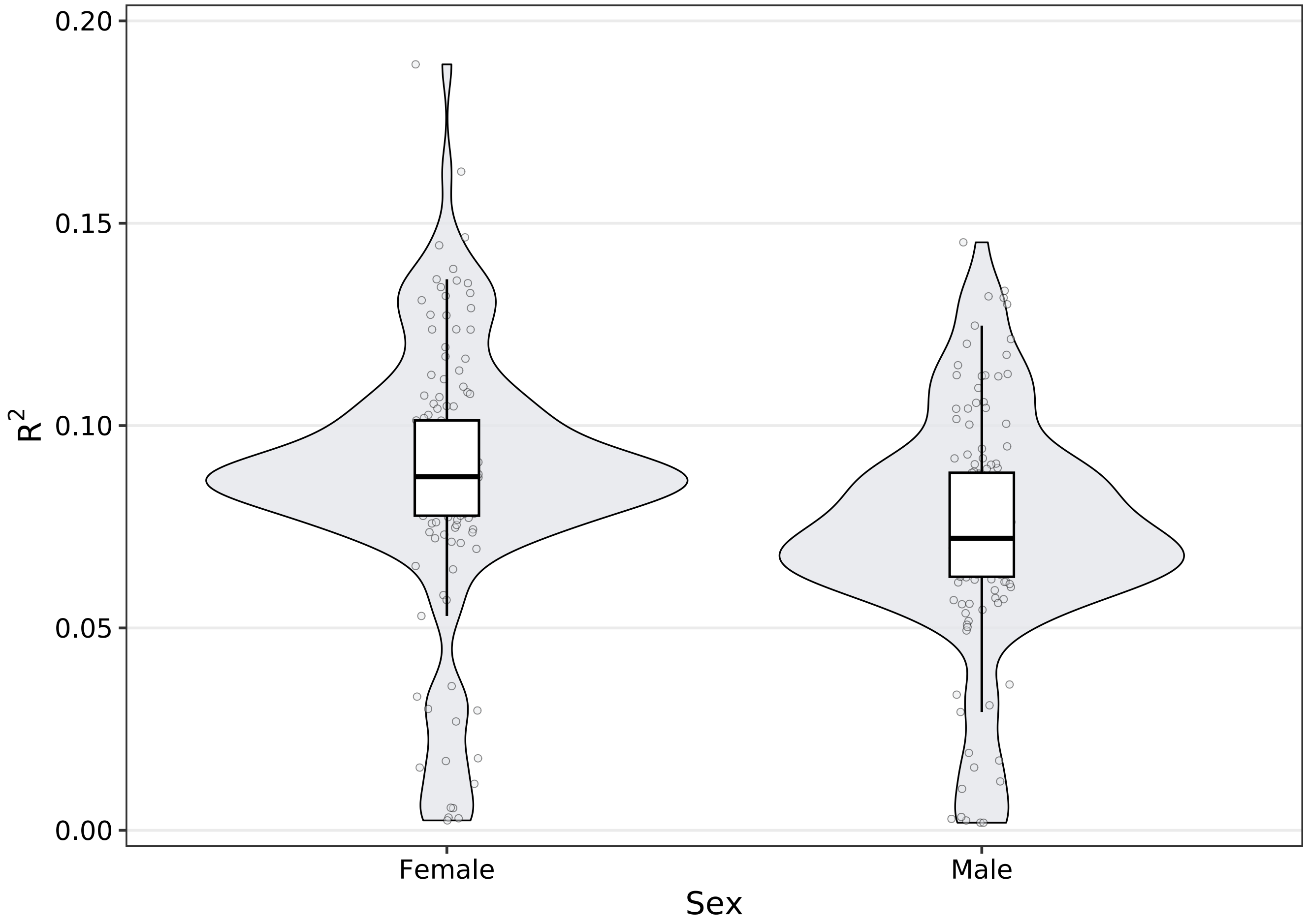


The violin plots and overlaid boxplots show the distribution of $R^2$ values for 141 metabolomic traits predicted by Mexico City Prospective Study (MCPS)-trained models in the 20% of MCPS participants withheld for internal validation, stratified by sex. The boxplots show the median, interquartile range, 1.5 times the interquartile range, and outliers; points show the values for individual metabolomic traits. Performance was better among female participants (median $R^2$: 0.087; IQR: 0.024) than among male participants (median $R^2$: 0.072; IQR: 0.026).

**Supplementary Fig. 7: Predictive performance of MCPS-trained models in the withheld MCPS subset by age group**

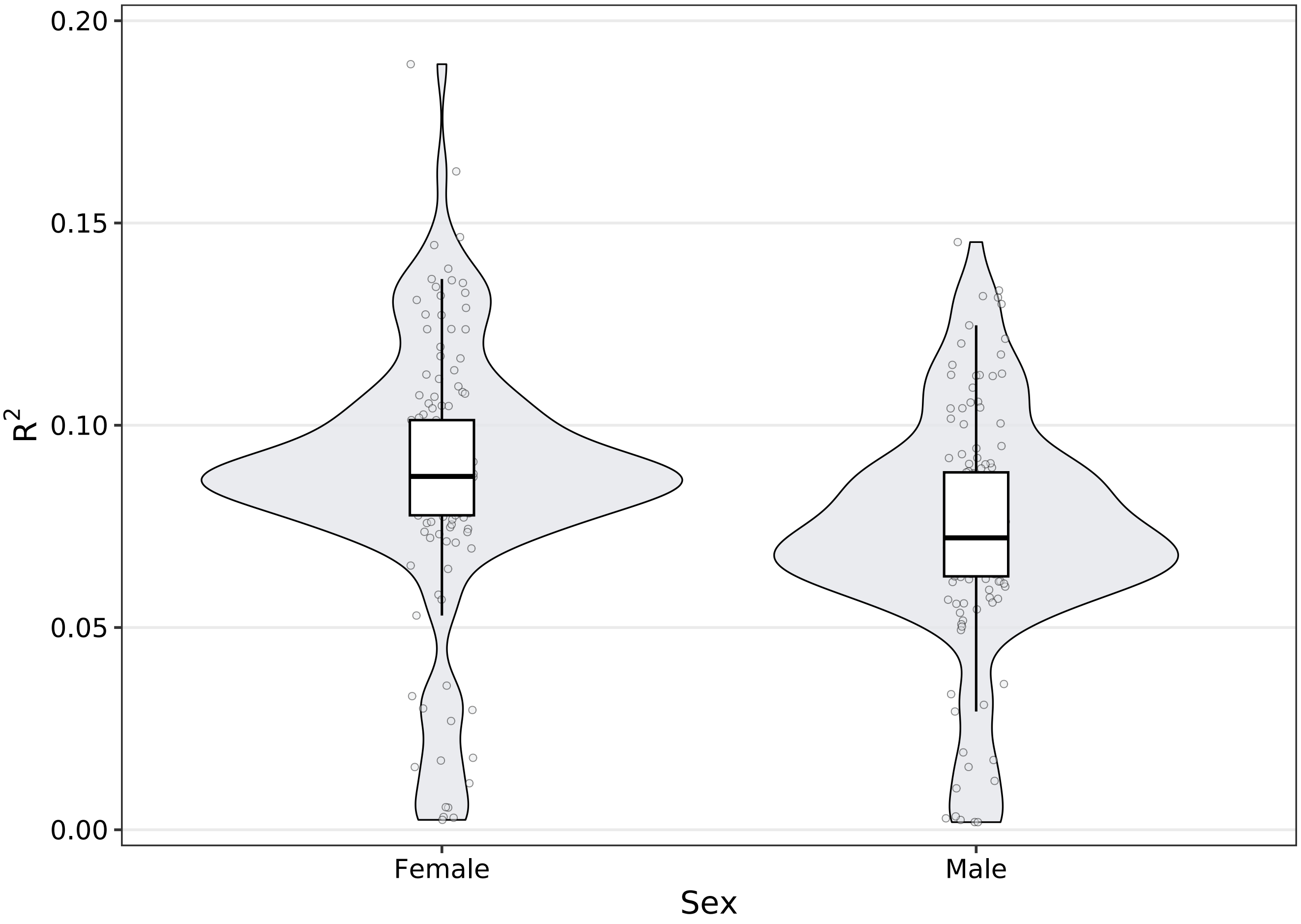


The violin plots and overlaid boxplots show the distribution of $R^2$ values for 141 metabolomic traits predicted by Mexico City Prospective Study (MCPS)-trained models in the 20% of MCPS participants withheld for internal validation, stratified by age at baseline. Predictive performance was higher among participants aged 35–49 years (median $R^2$: 0.084; IQR: 0.022) and 50–64 years (median $R^2$: 0.088; IQR: 0.022) than among participants aged ≥65 years (median $R^2$: 0.075; IQR: 0.032).

**Supplementary Fig. 8: Predictive performance of MCPS-trained models in the withheld MCPS subset by diabetes and baseline health status**

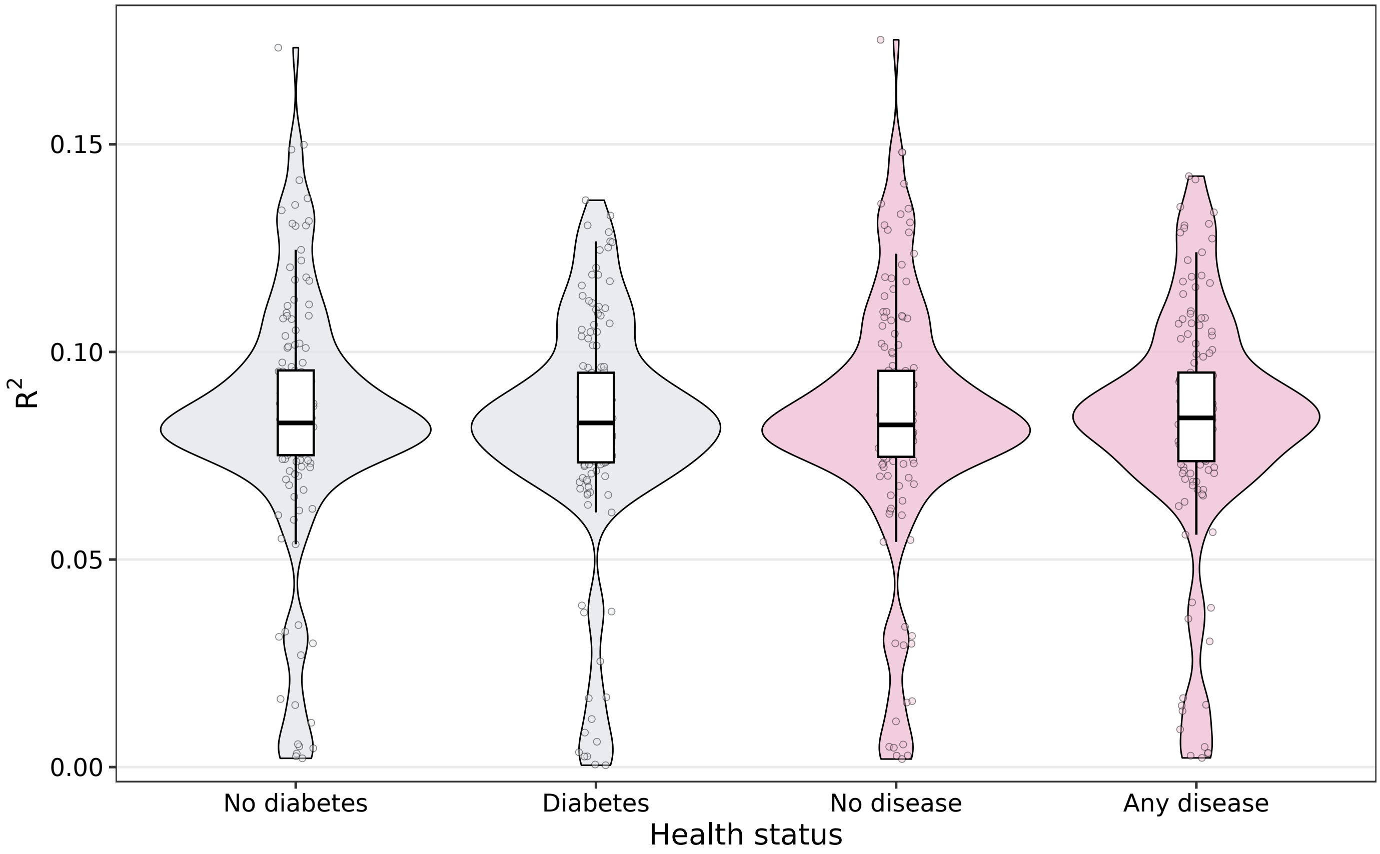


The violin plots and overlaid boxplots show the distribution of $R^2$ values for 141 metabolomic traits predicted by Mexico City Prospective Study (MCPS)-trained models in the 20% of MCPS participants withheld for internal validation, stratified by diabetes status and by the presence or absence of any of a range of common diseases at baseline. Median $R^2$ was 0.083 among participants without diabetes (IQR: 0.021) and 0.083 among participants with diabetes (IQR: 0.022). Median $R^2$ was 0.082 (IQR: 0.021) among participants with no disease and 0.084 (IQR: 0.021) among participants with at least one disease.

**Supplementary Fig. 9: Strength of statistical evidence of associations between cardiometabolic diseases and metabolomic traits predicted by MCPS- and INTERVAL-trained models in the AMR-ancestries subset of AoU**

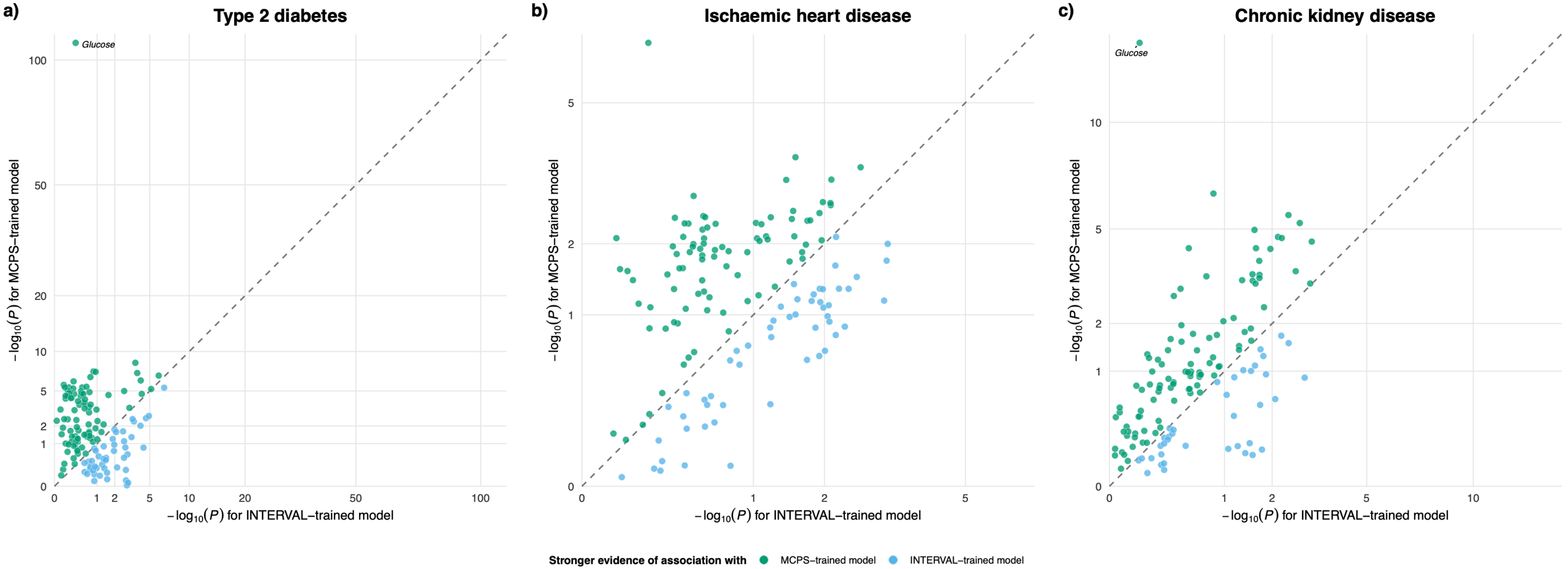


The statistical evidence for the estimated associations of 139 metabolomic traits, predicted using two different sets of models, with three high-burden cardiometabolic diseases: **a**, type 2 diabetes; **b**, ischaemic heart disease; and **c**, chronic kidney disease. For each metabolomic trait, the $-\log_{10}(P)$ values for the associations estimated using the MCPS- and INTERVAL-trained models are plotted on the y and the x axes, respetively. Associations were estimated using logistic regression, adjusting for age at the last electronic health record event, sex, and the first 10 genetic principal components. Point colours indicate whether stronger evidence of association was obtained using the MCPS-trained model (green) or the INTERVAL-trained model (blue). The dashed line represents the identity line, with points above the line indicating stronger evidence using the MCPS-trained model. Stronger evidence was obtained using MCPS-trained models for 92 of 139 traits for type 2 diabetes, 86 of 139 traits for ischaemic heart disease, and 101 of 139 traits for chronic kidney disease. The corresponding numbers of false discovery rate (FDR-)significant associations obtained using MCPS- compared to INTERVAL-trained models were 69 vs. 27, 55 vs. 3, and 26 vs. 0, respectively. *P* values shown on the axes are unadjusted; the FDR correction was applied separately within each disease and model set. Axes use square-root spacing to accommodate the wide range of *P* values.

### Supplementary Fig. 10: Associations of metabolomic traits predicted by MCPS- and INTERVAL-trained models with type 2 diabetes in the AMR-ancestries subset of AoU

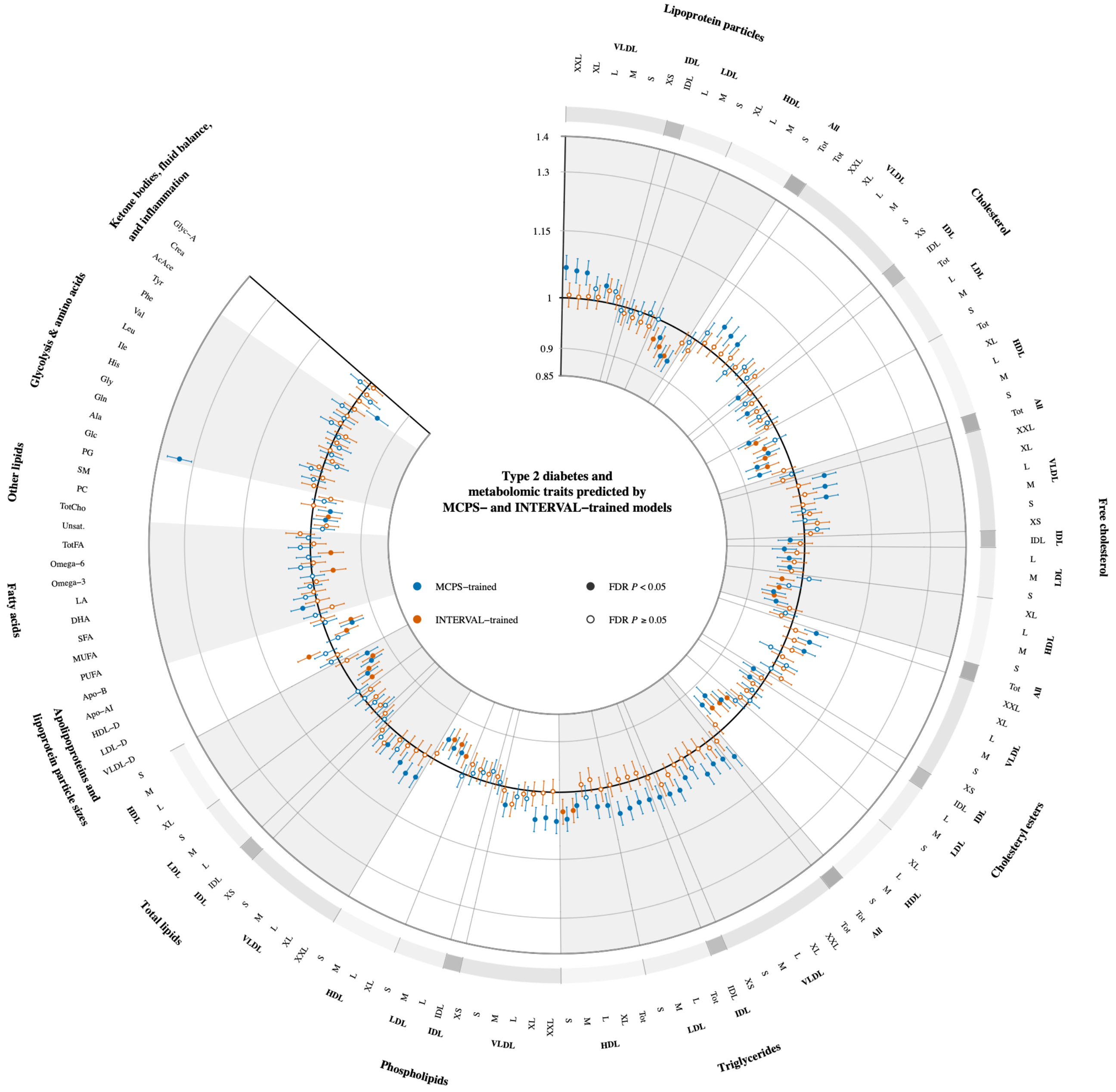


The wheel plot shows odds ratios for associations of 139 metabolomic traits predicted by Mexico City Prospective Study (MCPS)-trained and INTERVAL-trained models with type 2 diabetes in the subset of the All of Us Research Program (AoU) of admixed American (AMR) genetic ancestries. Odds ratios and 95% confidence intervals are shown per 1-s.d. higher predicted metabolomic trait. Blue points show estimates based on MCPS-trained models and orange points show estimates based on INTERVAL-trained models; filled points indicate associations with FDR-corrected *P* < 0.05 and open points indicate associations with FDR-corrected *P* ≥ 0.05. Grey sectors indicate metabolomic trait classes, and the outer band and radial guide lines distinguish VLDL, IDL, LDL, and HDL subclasses within lipoprotein-related classes.

Abbreviations: AoU: All of Us Research Program; AMR: admixed American; FDR: false discovery rate; s.d.: standard deviation; CI: confidence interval; XXL: chylomicrons and extremely large VLDL; XL: very large; L: large; M: medium; S: small; XS: very small; VLDL: very-low-density lipoprotein; IDL: intermediate-density lipoprotein; LDL: low-density lipoprotein; HDL: high-density lipoprotein; VLDL-D: average diameter for VLDL particles; LDL-D: average diameter for LDL particles; HDL-D: average diameter for LDL particles; Apo-AI: apolipoprotein A1; Apo-B: apolipoprotein B; Tot-C: total cholesterol; VLDL-C: VLDL cholesterol; LDL-C: LDL cholesterol; HDL-C: HDL cholesterol; Tot-FC: total free cholesterol; Tot-CE: total esterified cholesterol; Tot-TG: total triglycerides; VLDL-TG: triglycerides in VLDL; LDL-TG: triglycerides in LDL; HDL-TG: triglycerides in HDL; PUFA: polyunsaturated fatty acids; MUFA: monounsaturated fatty acids; SFA: saturated fatty acids; DHA: docosahexaenoic acid; LA: linoleic acid; Omega-3: omega-3 fatty acids; Omega-6: omega-6 fatty acids; TotFA: total fatty acids; Unsat.: degree of unsaturation; TotCho: total cholines; PC: phosphatidylcholines; SM: sphingomyelins; PG: phosphoglycerides; Glc: glucose; Ala: alanine; Gln: glutamine; Gly: glycine; His: histidine; Ile: isoleucine; Leu: leucine; Val: valine; Phe: phenylalanine; Tyr: tyrosine; AcAce: acetoacetate; Crea: creatinine; Glyc-A: glycoprotein acetyls.

### Supplementary Fig. 11: Associations of metabolomic traits predicted by MCPS- and INTERVAL-trained models with ischaemic heart disease in the AMR-ancestries subset of AoU

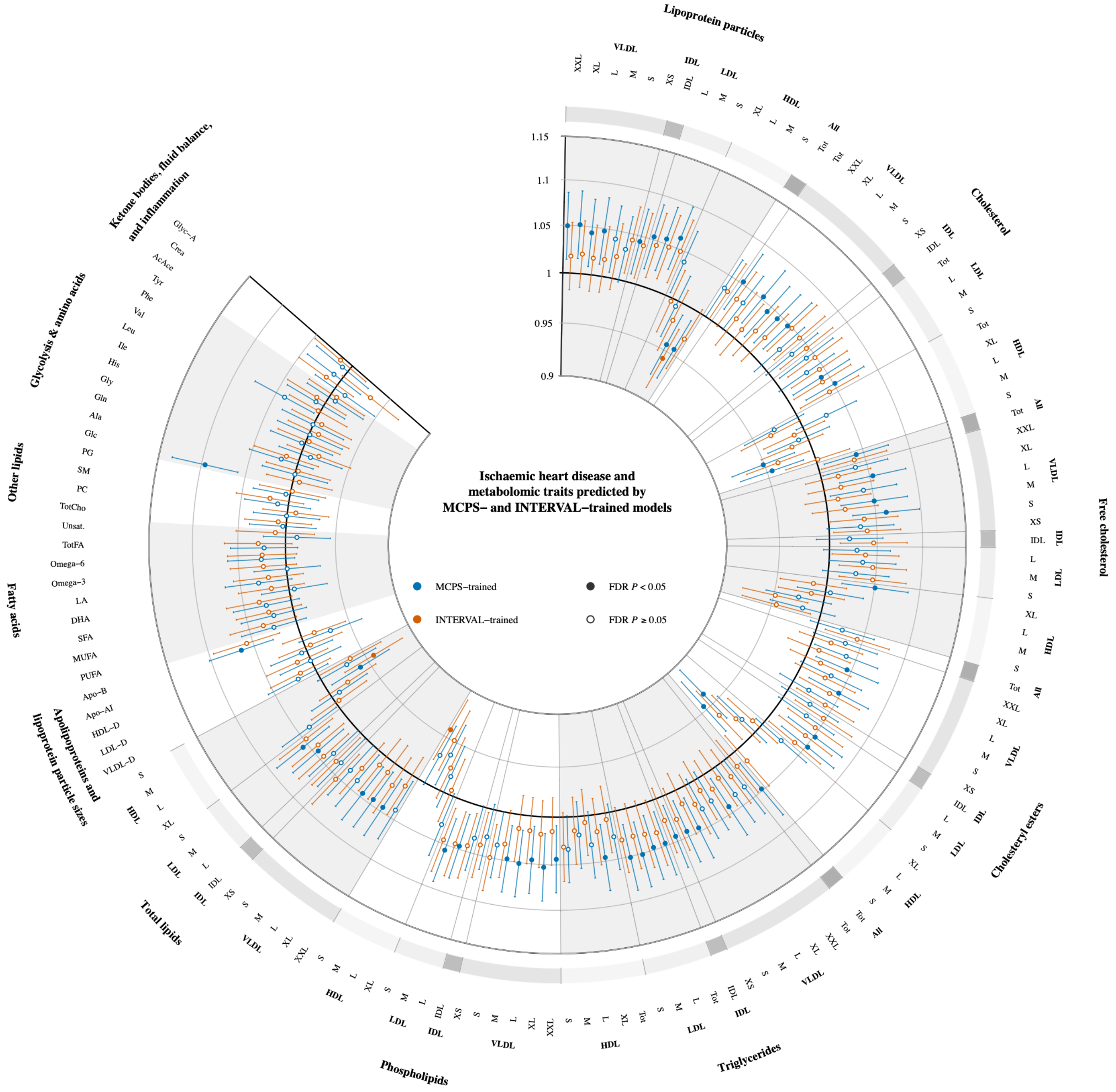


The wheel plot shows odds ratios for associations of 139 metabolomic traits predicted by Mexico City Prospective Study (MCPS)-trained and INTERVAL-trained models with ischaemic heart disease in the subset of the All of Us Research Program (AoU) of admixed American (AMR) genetic ancestries. Odds ratios and 95% confidence intervals are shown per 1-s.d. higher predicted metabolomic trait. Blue points show estimates based on MCPS-trained models and orange points show estimates based on INTERVAL-trained models; filled points indicate associations with FDR-corrected $P < 0.05$ and open points indicate associations with FDR-corrected $P \geq 0.05$. Grey sectors indicate metabolomic trait classes, and the outer band and radial guide lines distinguish VLDL, IDL, LDL, and HDL subclasses within lipoprotein-related classes.

Abbreviations: AoU: All of Us Research Program; AMR: admixed American; FDR: false discovery rate; s.d.: standard deviation; CI: confidence interval; XXL: chylomicrons and extremely large VLDL; XL: very large; L: large; M: medium; S: small; XS: very small; VLDL: very-low-density lipoprotein; IDL: intermediate-density lipoprotein; LDL: low-density lipoprotein; HDL: high-density lipoprotein; VLDL-D: average diameter for VLDL particles; LDL-D: average diameter for LDL particles; HDL-D: average diameter for LDL particles; Apo-AI: apolipoprotein A1; Apo-B: apolipoprotein B; Tot-C: total cholesterol; VLDL-C: VLDL cholesterol; LDL-C: LDL cholesterol; HDL-C: HDL cholesterol; Tot-FC: total free cholesterol; Tot-CE: total esterified cholesterol; Tot-TG: total triglycerides; VLDL-TG: triglycerides in VLDL; LDL-TG: triglycerides in LDL; HDL-TG: triglycerides in HDL; PUFA: polyunsaturated fatty acids; MUFA: monounsaturated fatty acids; SFA: saturated fatty acids; DHA: docosahexaenoic acid; LA: linoleic acid; Omega-3: omega-3 fatty acids; Omega-6: omega-6 fatty acids; TotFA: total fatty acids; Unsat.: degree of unsaturation; TotCho: total cholines; PC: phosphatidylcholines; SM: sphingomyelins; PG: phosphoglycerides; Glc: glucose; Ala: alanine; Gln: glutamine; Gly: glycine; His: histidine; Ile: isoleucine; Leu: leucine; Val: valine; Phe: phenylalanine; Tyr: tyrosine; AcAce: acetoacetate; Crea: creatinine; Glyc-A: glycoprotein acetyls.

**Supplementary Fig. 12: Associations of metabolomic traits predicted by MCPS- and INTERVAL-trained models with chronic kidney disease in the AMR-ancestries subset of AoU**

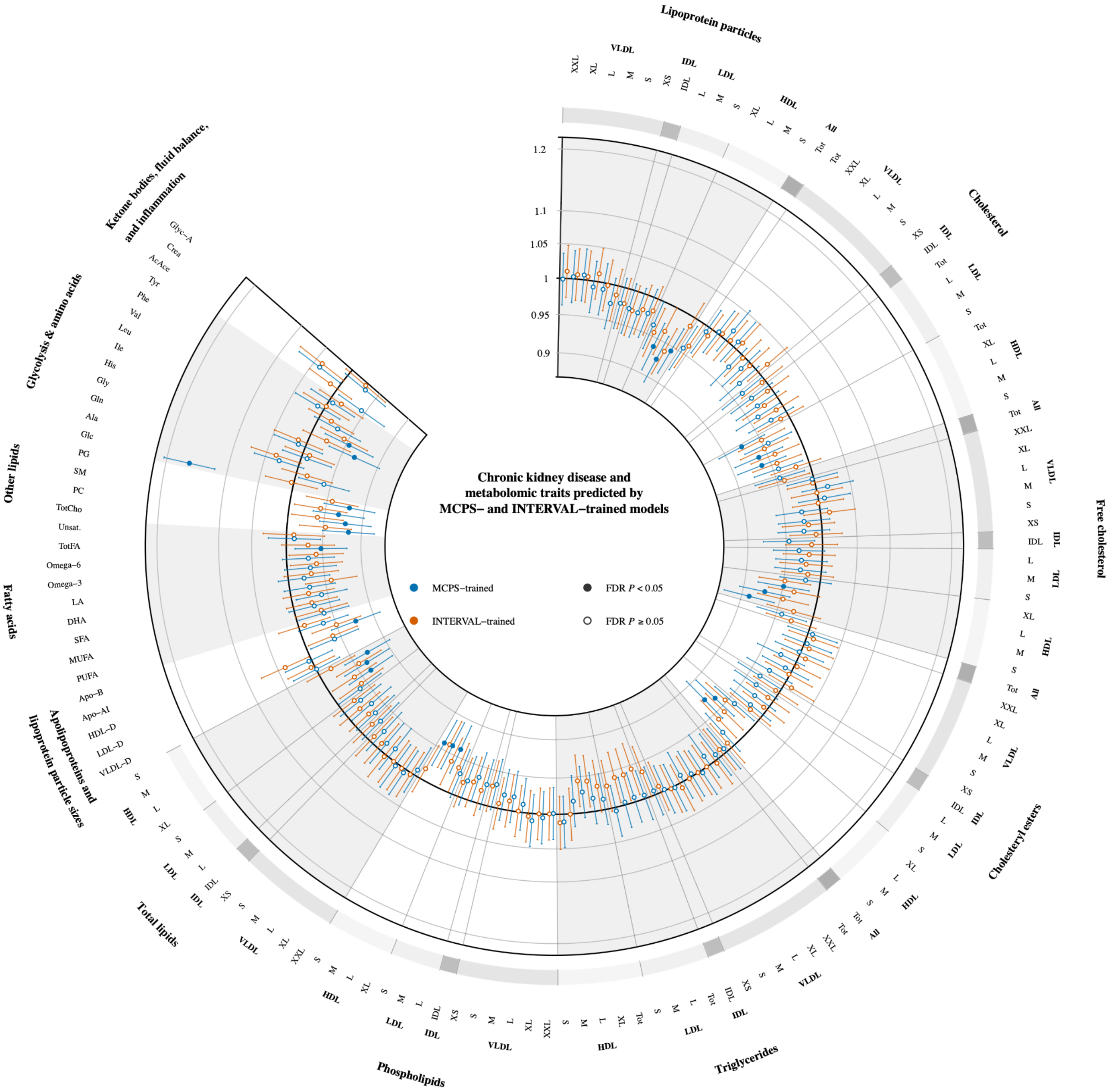


The wheel plot shows odds ratios for associations of 139 metabolomic traits predicted by Mexico City Prospective Study (MCPS)-trained and INTERVAL-trained models with chronic kidney disease in the subset of the All of Us Research Program (AoU) of admixed American (AMR) genetic ancestries. Odds ratios and 95% confidence intervals are shown per 1-s.d. higher predicted metabolomic trait. Blue points show estimates based on MCPS-trained models and orange points show estimates based on INTERVAL-trained models; filled points indicate associations with FDR-corrected $P < 0.05$ and open points indicate associations with FDR-corrected $P \geq 0.05$. Grey sectors indicate metabolomic trait classes, and the outer band and radial guide lines distinguish VLDL, IDL, LDL, and HDL subclasses within lipoprotein-related classes.

Abbreviations: AoU: All of Us Research Program; AMR: admixed American; FDR: false discovery rate; s.d.: standard deviation; CI: confidence interval; XXL: chylomicrons and extremely large (VLDL); XL: very large; L: large; M: medium; S: small; XS: very small; VLDL: very-low-density lipoprotein; IDL: intermediate-density lipoprotein; LDL: low-density lipoprotein; HDL: high-density lipoprotein; Tot: total; VLDL-D: average diameter for VLDL particles; LDL-D: average diameter for LDL particles; HDL-D: average diameter for LDL particles; Apo-AI: apolipoprotein A1; Apo-B: apolipoprotein B; Tot-C: total cholesterol; VLDL-C: VLDL cholesterol; LDL-C: LDL cholesterol; HDL-C: HDL cholesterol; Tot-FC: total free cholesterol; Tot-CE: total esterified cholesterol; Tot-TG: total triglycerides; VLDL-TG: triglycerides in VLDL; LDL-TG: triglycerides in LDL; HDL-TG: triglycerides in HDL; PUFA: polyunsaturated fatty acids; MUFA: monounsaturated fatty acids; SFA: saturated fatty acids; DHA: docosahexaenoic acid; LA: linoleic acid; Omega-3: omega-3 fatty acids; Omega-6: omega-6 fatty acids; TotFA: total fatty acids; Unsat.: degree of unsaturation; TotCho: total cholines; PC: phosphatidylcholines; SM: sphingomyelins; PG: phosphoglycerides; Glc: glucose; Ala: alanine; Gln: glutamine; Gly: glycine; His: histidine; Ile: isoleucine; Leu: leucine; Val: valine; Phe: phenylalanine; Tyr: tyrosine; AcAce: acetoacetate; Crea: creatinine; Glyc-A: glycoprotein acetyls.
