## Supplementary note 1 for "Enhanced power and transferability for genetics-driven metabolomic biomarker discovery in admixed American cohorts"

### Details of genome wide association studies of metabolomic traits conducted prior to the present study in the Mexico City Prospective Study

Genome-wide association studies (GWAS) of baseline Nightingale Health nuclear magnetic resonance (NMR) metabolomic traits in the Mexico City Prospective Study (MCPS) were conducted as part of a broader, comprehensive GWAS Atlas project for phenotypes of various categories in MCPS and shared with the authors of the present work via private communication. The present work used the summary statistics for the 141 directly measured NMR metabolomic traits. Trait-specific GWAS samples comprised 123,022 to 130,841 participants with linked genotype and phenotype data and non-missing metabolomic values. Participants with a possible X/Y chromosome aneuploidy or unreliable genotype-phenotype linkage were excluded from all GWAS. For NMR traits, participants of ages > 79 years or taking lipid-lowering medication were additionally excluded. Zero trait values were replaced before analysis by a random value between 0 and the lowest observed non-zero value for the corresponding trait.

Association testing was performed with REGENIE v4.0[1]. In step 1, whole-genome regression models were fitted using directly genotyped variants filtered to minor allele count (MAC) ≥ 100, with leave-one-chromosome-out predictions carried forward to step 2. In step 2, association tests were run using TOPMed-imputed genotype dosages in BGEN format[2]. Variants were restricted to those passing imputation-quality filtering (imputation INFO $r^2 > 0.4$) and a minimum MAC of 1. NMR traits were analysed as continuous phenotypes, and rank-based inverse normal transformation was applied in REGENIE (--apply-rint). Models included age, age squared, sex, district of residence and the first seven genetic principal components, and quintiles of fasting duration.

Chromosome-specific REGENIE outputs were combined into phenotype-specific summary statistics. Variant quality control (QC) for downstream use was based on the informative sample for each trait: allele frequencies were estimated within the trait-specific informative sample, and effective sample size was calculated using allele frequency, trait-specific informative $N$ and imputation INFO, with sex-specific allele frequencies and sample sizes used for chromosome X. Variants with effective sample size < 30 were excluded. Genome-wide significant associations used for the model-training variant selection described in the main text were defined as those having $P < 5 \times 10^{-8}$.

1. Mbatchou J, Barnard L, Backman J, et al. Computationally efficient whole-genome regression for quantitative and binary traits. *Nature Genetics*. 2021/07/01 2021;53(7):1097-1103. doi:10.1038/s41588-021-00870-7
2. Taliun D, Harris DN, Kessler MD, et al. Sequencing of 53,831 diverse genomes from the NHLBI TOPMed Program. *Nature*. 2021/02/01 2021;590(7845):290-299. doi:10.1038/s41586-021-03205-y
